# Augmenting mortality prediction with medication data and machine learning models

**DOI:** 10.1101/2024.04.16.24305420

**Authors:** Brian Murray, Tianyi Zhang, Amoreena Most, Xianyan Chen, Susan E. Smith, John W. Devlin, David J. Murphy, Andrea Sikora, Rishikesan Kamaleswaran, the MRC-ICU Investigator Team

## Abstract

**Background:** In critically ill patients, complex relationships exist among patient disease factors, medication management, and mortality. Considering the potential for nonlinear relationships and the high dimensionality of medication data, machine learning and advanced regression methods may offer advantages over traditional regression techniques. The purpose of this study was to evaluate the role of different modeling approaches incorporating medication data for mortality prediction.

**Methods:** This was a single-center, observational cohort study of critically ill adults. A random sample of 991 adults admitted ≥ 24 hours to the intensive care unit (ICU) from 10/2015 to 10/2020 were included. Models to predict hospital mortality at discharge were created. Models were externally validated against a temporally separate dataset of 4,878 patients. Potential mortality predictor variables (n=27, together with 14 indicators for missingness) were collected at baseline (age, sex, service, diagnosis) and 24 hours (illness severity, supportive care use, fluid balance, laboratory values, MRC-ICU score, and vasopressor use) and included in all models. The optimal traditional (equipped with linear predictors) logistic regression model and optimal advanced (equipped with nature splines, smoothing splines, and local linearity) logistic regression models were created using stepwise selection by Bayesian information criterion (BIC). Supervised, classification-based ML models [e.g., Random Forest, Support Vector Machine (SVM), and XGBoost] were developed. Area under the receiver operating characteristic (AUROC), positive predictive value (PPV), and negative predictive value (NPV) were compared among different mortality prediction models.

**Results:** A model including MRC-ICU in addition to SOFA and APACHE II demonstrated an AUROC of 0.83 for hospital mortality prediction, compared to AUROCs of 0.72 and 0.81 for APACHE II and SOFA alone. Machine learning models based on Random Forest, SVM, and XGBoost demonstrated AUROCs of 0.83, 0.85, and 0.82, respectively. Accuracy of traditional regression models was similar to that of machine learning models. MRC-ICU demonstrated a moderate level of feature importance in both XGBoost and Random Forest. Across all ten models, performance was lower on the validation set.

**Conclusions:** While medication data were not included as a significant predictor in regression models, addition of MRC-ICU to severity of illness scores (APACHE II and SOFA) improved AUROC for mortality prediction. Machine learning methods did not improve model performance relative to traditional regression methods.

## Introduction

Medications are key influencers of mortality but are rarely included in mortality prediction models. Assessing medications as direct determinants of mortality is difficult given complex interactions with patient physiology, disease pathophysiology, and other elements of a medication regimen. Moreover, the therapeutic effect of any medication is balanced against the potential to cause harm.[1] Patients in the intensive care unit (ICU) are particularly susceptible to the effects of medications as they receive on average twice as many medications as patients on a general ward,[2, 3] are frequently exposed to clinically relevant potential drug-drug interactions (DDIs),[4, 5] and are more than twice as likely to experience an adverse drug event (ADE).[2, 3, 6–8] Elucidating the direct effects of medications on mortality of ICU patients and incorporating that information into prediction models may improve model performance and better inform care of ICU patients.

The incorporation of medication data into prediction models for critically ill patients has improved results for prediction of fluid overload and prolonged duration of mechanical ventilation, particularly with the application of supervised machine learning approaches.[9, 10] Additionally, a pilot study of six machine learning methods also showed that incorporation of medication data and the medication regimen complexity-intensive care unit (MRC-ICU) score improved mortality prediction, and adding MRC-ICU to severity of illness improved traditional regression as well.[11] These examples offer credence to the concept that incorporating information on medication regimens is useful in predicting both shot-term and long-term outcomes for ICU patients.

Development of accurate mortality prediction models remains an ongoing area of interest in critically ill patients.[12–14] Severity of illness scores (Acute Physiology and Chronic Health Evaluation [APACHE]-IV, Sequential Organ Failure Assessment [SOFA], and Simplified Acute Physiology Score [SAPS] I-III) are commonly used to predict mortality in the ICU but have potential limitations.[15, 16] While these scores are reflective of syndrome-attributable mortality, they do not consider the impact of patient management and the contribution of clinical decision-making to patient outcomes.

Artificial intelligence (AI) and machine learning have renewed interest in outcome prediction in a variety of patient populations, including the critically ill. The ability of AI algorithms to process vast amounts of information and identify previously unelucidated patterns in data highlight the potential for these methodologies to improve prediction models through the incorporation of a greater number of latent outcome determinants and markers with predictive value. Using machine learning, it is possible to incorporate specific and granular medication data with severity of illness variables to create more comprehensive outcome prediction models. The purpose of this study was to assess the value of medication data in the context of other patient specific factors (e.g., severity of illness) in machine learning-based mortality prediction. We hypothesized that incorporation of medication data would significantly improve model performance and that medications would be highly ranked in feature importance graphs.

## Methods

### Study Population

This retrospective, observational study was reviewed by the University of Georgia (UGA) Institutional Review Board (IRB) and deemed to be exempt from IRB oversight (Project00001541). All methods were performed in accordance with the ethical standards of the UGA IRB and the Helinski Declaration of 1975. Patient data were obtained via the Carolina Data Warehouse, which houses EPIC^®^ electronic health record (EHR) data from the University of North Carolina Health System (UNCHS), an integrated healthcare delivery system. Given the intensity of the data collection effort, we employed random number generation to identify a sample of 1,000 adults aged ≥ 18 years admitted to the ICU ≥ 24 hours between October 2015 and October 2020. Only data from the first ICU admission for each patient were included. Patients were excluded if they were placed on comfort care within the first 24 hours of their ICU stay. These methods have been previously published.[9, 17] This evaluation followed the STROBE (Strengthening the Reporting of Observational Studies in Epidemiology) and CONSORT-AI (Consolidated Standards of Reporting Trials-Artificial Intelligence) extension reporting frameworks, as applicable.[18, 19]

### Data Collection and Outcomes

The primary outcome was hospital mortality. The EHR was queried to obtain: 1) *ICU baseline characteristics:* age, sex, type of admission ICU (i.e., burn, cardiac, cardiothoracic, medical, neurosciences, surgical, mixed), primary ICU admission diagnosis (i.e., burn, cardiovascular, dermatology, electrolyte abnormalities, endocrine, fever, gastrointestinal, hematologic, hepatic, infection, mental health, neoplasm, neurology, pneumonia, pregnancy, pulmonary, renal, respiratory, sepsis, shock, syncope, toxicology/ingestion, trauma, weakness, or other); 2) *Clinical data 24 hours after ICU admission:* Acute Physiology and Chronic Health Evaluation (APACHE) II and Sequential Organ Failure Assessment (SOFA) score (using worst values in the 24 hour period), vital signs (i.e., heart rate, systolic blood pressure, temperature), Acute Respiratory Distress Syndrome (ARDS) classification (i.e., mild, moderate, or severe based on ratio of partial pressure of arterial oxygen [PaO_2_] to fraction of inspire oxygen [FiO_2_]), use of supportive care devices (i.e., renal replacement therapy, mechanical ventilation), serum laboratory values (i.e., albumin, bicarbonate, creatinine, glucose, lactate, potassium, pH, sodium, hemoglobin, hematocrit, platelets, white blood cell count), fluid balance (mL); 3) *Medication data at 24 hour:* MRC-ICU score and vasopressor use within 24 hour.

### Data Analysis

**Missingness:** For missing components of SOFA and APACHE II score, patients were considered to have ‘normal’ condition for that corresponding variable (Supplemental Digital Content – Table 1). Additionally, Dummy Variable Adjustment for missingness was conducted for 14 additional predictors to assess how much the missing cases differed from an average individual without missing data for the primary outcome of mortality.

**Table 1.**
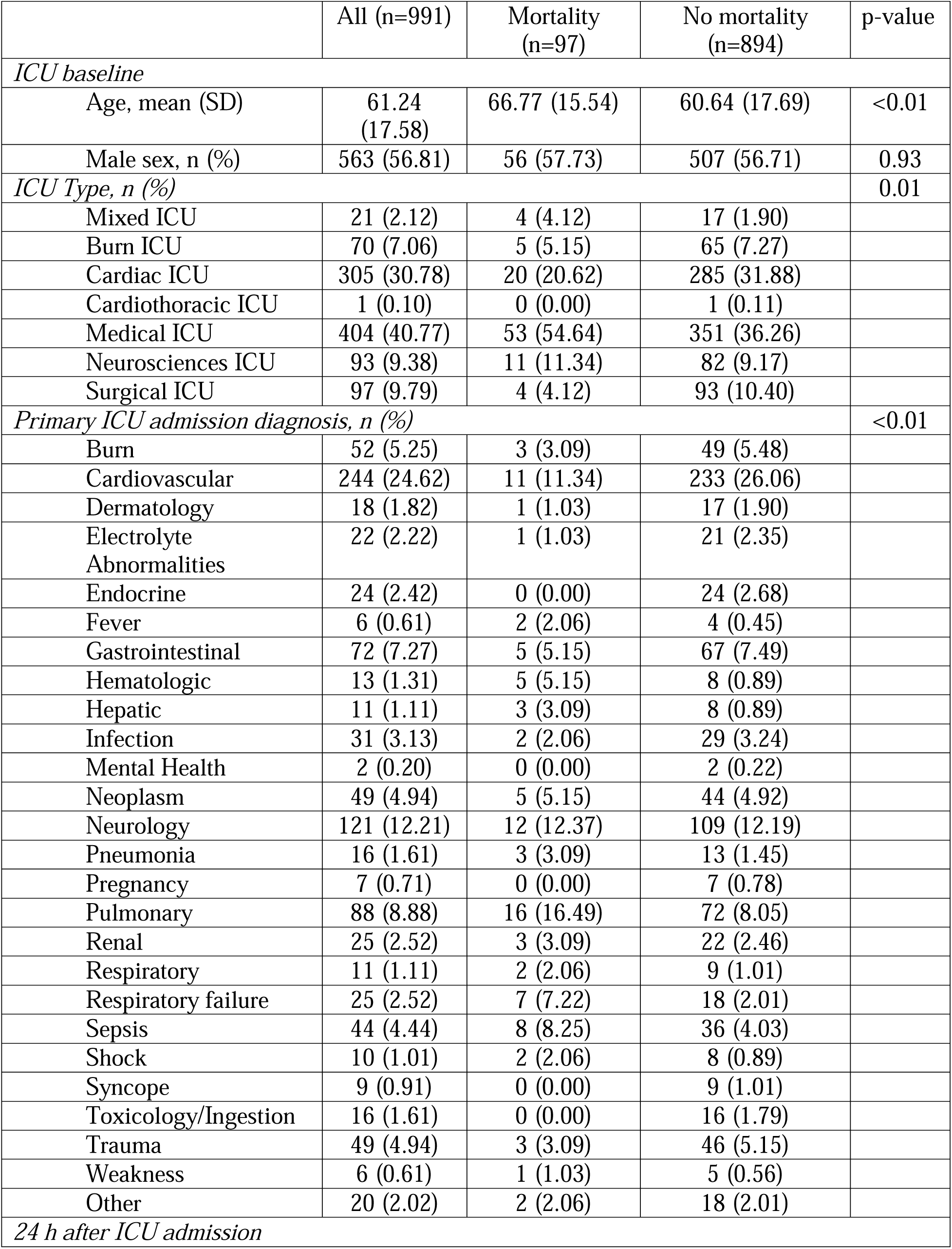
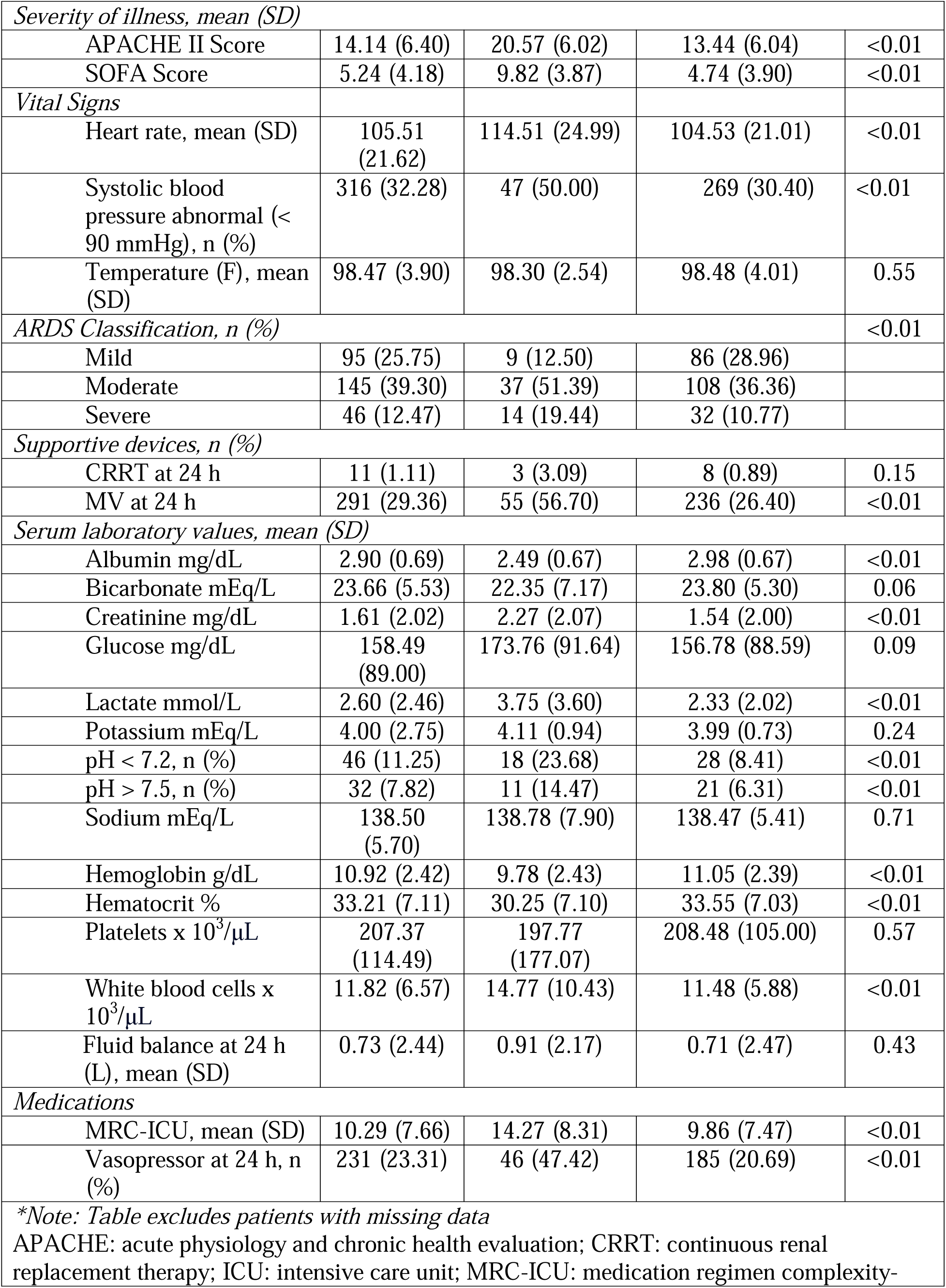
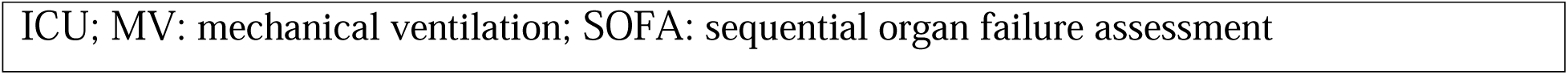
Baseline study cohort demographics.

|  | All (n=991) | Mortality<br>(n=97) | No mortality<br>(n=894) | p-value |
| --- | --- | --- | --- | --- |
| *ICU baseline* |  |  |  |  |
| Age, mean (SD) | 61.24<br>(17.58) | 66.77 (15.54) | 60.64 (17.69) | <0.01 |
| Male sex, n (%) | 563 (56.81) | 56 (57.73) | 507 (56.71) | 0.93 |
| *ICU Type, n (%)* |  |  |  | 0.01 |
| Mixed ICU | 21 (2.12) | 4 (4.12) | 17 (1.90) |  |
| Burn ICU | 70 (7.06) | 5 (5.15) | 65 (7.27) |  |
| Cardiac ICU | 305 (30.78) | 20 (20.62) | 285 (31.88) |  |
| Cardiothoracic ICU | 1 (0.10) | 0 (0.00) | 1 (0.11) |  |
| Medical ICU | 404 (40.77) | 53 (54.64) | 351 (36.26) |  |
| Neurosciences ICU | 93 (9.38) | 11 (11.34) | 82 (9.17) |  |
| Surgical ICU | 97 (9.79) | 4 (4.12) | 93 (10.40) |  |
| *Primary ICU admission diagnosis, n (%)* |  |  |  | <0.01 |
| Burn | 52 (5.25) | 3 (3.09) | 49 (5.48) |  |
| Cardiovascular | 244 (24.62) | 11 (11.34) | 233 (26.06) |  |
| Dermatology | 18 (1.82) | 1 (1.03) | 17 (1.90) |  |
| Electrolyte<br>Abnormalities | 22 (2.22) | 1 (1.03) | 21 (2.35) |  |
| Endocrine | 24 (2.42) | 0 (0.00) | 24 (2.68) |  |
| Fever | 6 (0.61) | 2 (2.06) | 4 (0.45) |  |
| Gastrointestinal | 72 (7.27) | 5 (5.15) | 67 (7.49) |  |
| Hematologic | 13 (1.31) | 5 (5.15) | 8 (0.89) |  |
| Hepatic | 11 (1.11) | 3 (3.09) | 8 (0.89) |  |
| Infection | 31 (3.13) | 2 (2.06) | 29 (3.24) |  |
| Mental Health | 2 (0.20) | 0 (0.00) | 2 (0.22) |  |
| Neoplasm | 49 (4.94) | 5 (5.15) | 44 (4.92) |  |
| Neurology | 121 (12.21) | 12 (12.37) | 109 (12.19) |  |
| Pneumonia | 16 (1.61) | 3 (3.09) | 13 (1.45) |  |
| Pregnancy | 7 (0.71) | 0 (0.00) | 7 (0.78) |  |
| Pulmonary | 88 (8.88) | 16 (16.49) | 72 (8.05) |  |
| Renal | 25 (2.52) | 3 (3.09) | 22 (2.46) |  |
| Respiratory | 11 (1.11) | 2 (2.06) | 9 (1.01) |  |
| Respiratory failure | 25 (2.52) | 7 (7.22) | 18 (2.01) |  |
| Sepsis | 44 (4.44) | 8 (8.25) | 36 (4.03) |  |
| Shock | 10 (1.01) | 2 (2.06) | 8 (0.89) |  |
| Syncope | 9 (0.91) | 0 (0.00) | 9 (1.01) |  |
| Toxicology/Ingestion | 16 (1.61) | 0 (0.00) | 16 (1.79) |  |
| Trauma | 49 (4.94) | 3 (3.09) | 46 (5.15) |  |
| Weakness | 6 (0.61) | 1 (1.03) | 5 (0.56) |  |
| Other | 20 (2.02) | 2 (2.06) | 18 (2.01) |  |
| *24 h after ICU admission* |  |  |  |  |
| *Severity of illness, mean (SD)* |  |  |  |  |
| APACHE II Score | 14.14 (6.40) | 20.57 (6.02) | 13.44 (6.04) | <0.01 |
| SOFA Score | 5.24 (4.18) | 9.82 (3.87) | 4.74 (3.90) | <0.01 |
| *Vital Signs* |  |  |  |  |
| Heart rate, mean (SD) | 105.51 (21.62) | 114.51 (24.99) | 104.53 (21.01) | <0.01 |
| Systolic blood pressure abnormal (< 90 mmHg), n (%) | 316 (32.28) | 47 (50.00) | 269 (30.40) | <0.01 |
| Temperature (F), mean (SD) | 98.47 (3.90) | 98.30 (2.54) | 98.48 (4.01) | 0.55 |
| *ARDS Classification, n (%)* |  |  |  | <0.01 |
| Mild | 95 (25.75) | 9 (12.50) | 86 (28.96) |  |
| Moderate | 145 (39.30) | 37 (51.39) | 108 (36.36) |  |
| Severe | 46 (12.47) | 14 (19.44) | 32 (10.77) |  |
| *Supportive devices, n (%)* |  |  |  |  |
| CRRT at 24 h | 11 (1.11) | 3 (3.09) | 8 (0.89) | 0.15 |
| MV at 24 h | 291 (29.36) | 55 (56.70) | 236 (26.40) | <0.01 |
| *Serum laboratory values, mean (SD)* |  |  |  |  |
| Albumin mg/dL | 2.90 (0.69) | 2.49 (0.67) | 2.98 (0.67) | <0.01 |
| Bicarbonate mEq/L | 23.66 (5.53) | 22.35 (7.17) | 23.80 (5.30) | 0.06 |
| Creatinine mg/dL | 1.61 (2.02) | 2.27 (2.07) | 1.54 (2.00) | <0.01 |
| Glucose mg/dL | 158.49 (89.00) | 173.76 (91.64) | 156.78 (88.59) | 0.09 |
| Lactate mmol/L | 2.60 (2.46) | 3.75 (3.60) | 2.33 (2.02) | <0.01 |
| Potassium mEq/L | 4.00 (2.75) | 4.11 (0.94) | 3.99 (0.73) | 0.24 |
| pH < 7.2, n (%) | 46 (11.25) | 18 (23.68) | 28 (8.41) | <0.01 |
| pH > 7.5, n (%) | 32 (7.82) | 11 (14.47) | 21 (6.31) | <0.01 |
| Sodium mEq/L | 138.50 (5.70) | 138.78 (7.90) | 138.47 (5.41) | 0.71 |
| Hemoglobin g/dL | 10.92 (2.42) | 9.78 (2.43) | 11.05 (2.39) | <0.01 |
| Hematocrit % | 33.21 (7.11) | 30.25 (7.10) | 33.55 (7.03) | <0.01 |
| Platelets x 10<sup>3</sup>/μL | 207.37 (114.49) | 197.77 (177.07) | 208.48 (105.00) | 0.57 |
| White blood cells x 10<sup>3</sup>/μL | 11.82 (6.57) | 14.77 (10.43) | 11.48 (5.88) | <0.01 |
| Fluid balance at 24 h (L), mean (SD) | 0.73 (2.44) | 0.91 (2.17) | 0.71 (2.47) | 0.43 |
| *Medications* |  |  |  |  |
| MRC-ICU, mean (SD) | 10.29 (7.66) | 14.27 (8.31) | 9.86 (7.47) | <0.01 |
| Vasopressor at 24 h, n (%) | 231 (23.31) | 46 (47.42) | 185 (20.69) | <0.01 |
| *\*Note: Table excludes patients with missing data*<br>APACHE: acute physiology and chronic health evaluation; CRRT: continuous renal replacement therapy; ICU: intensive care unit; MRC-ICU: medication regimen complexity- |  |  |  |  |

ICU; MV: mechanical ventilation; SOFA: sequential organ failure assessment

To evaluate the models, the cohort was split into training and test sets, using a ratio of 4:1. Four types of logistic regression models were created for predicting mortality with 27 variables and an additional 14 indicators for missingness (Supplemental Digital Content – Table 2) for both the training and test sets: 1) linear predictors, 2) predictors in nature cubic splines, 3) predictors in smoothing cubic splines, 4) local linear predictors. We then applied stepwise elimination with Bayesian Information Criterion (BIC) to select the final model.[20] Three machine learning models, Random Forest, SVM, and XGBoost, were employed.[21–26] Feature importance graphs were applied to visualize the strength of every predictor. For XGBoost, feature importance was measured as the frequency a feature was used in the trees. For Random Forest, feature importance was measured by mean decrease in node impurity. For SVM, feature importance was measured by the absolute magnitude of the coefficients for each variable with a normalized data set.

**Table 2.**
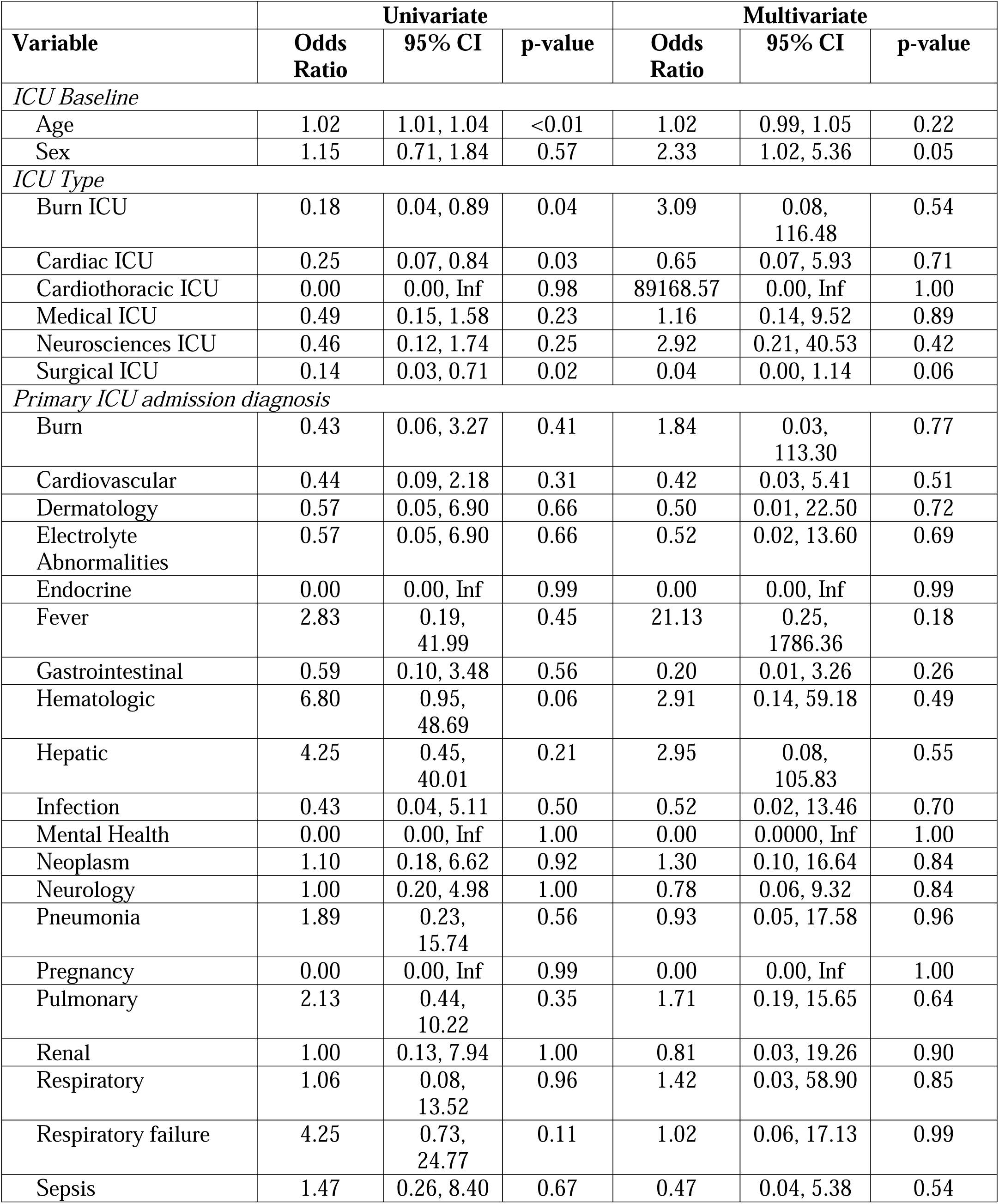
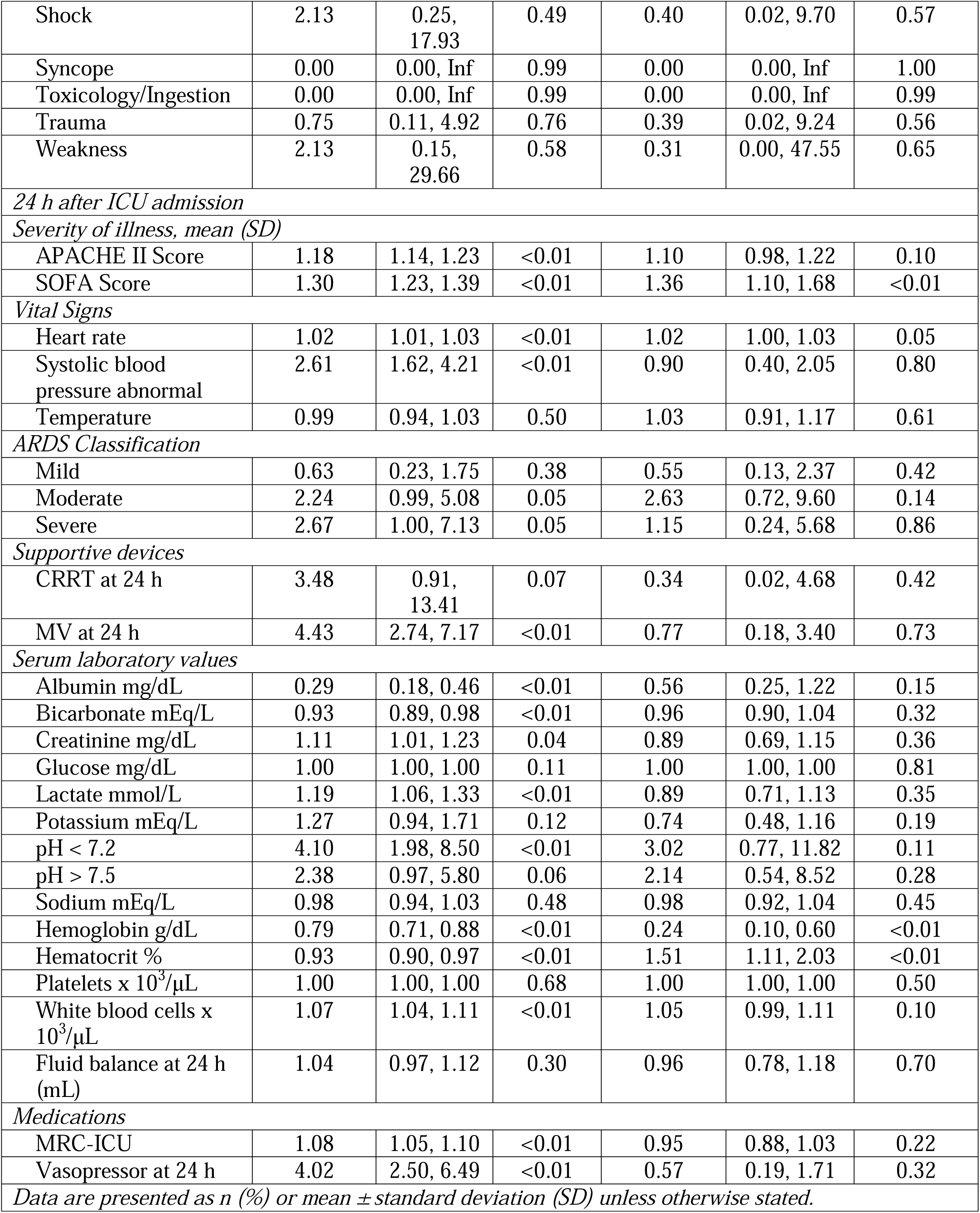
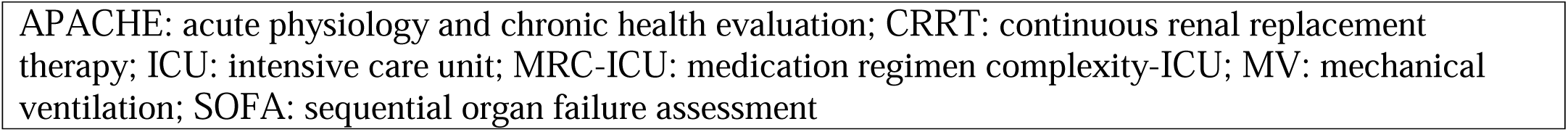
Mortality prediction variables univariate and multivariate analysis.

| Variable | Univariate |  |  | Multivariate |  |  |
| --- | --- | --- | --- | --- | --- | --- |
|  | Odds Ratio | 95% CI | p-value | Odds Ratio | 95% CI | p-value |
| *ICU Baseline* |  |  |  |  |  |  |
| Age | 1.02 | 1.01, 1.04 | <0.01 | 1.02 | 0.99, 1.05 | 0.22 |
| Sex | 1.15 | 0.71, 1.84 | 0.57 | 2.33 | 1.02, 5.36 | 0.05 |
| *ICU Type* |  |  |  |  |  |  |
| Burn ICU | 0.18 | 0.04, 0.89 | 0.04 | 3.09 | 0.08, 116.48 | 0.54 |
| Cardiac ICU | 0.25 | 0.07, 0.84 | 0.03 | 0.65 | 0.07, 5.93 | 0.71 |
| Cardiothoracic ICU | 0.00 | 0.00, Inf | 0.98 | 89168.57 | 0.00, Inf | 1.00 |
| Medical ICU | 0.49 | 0.15, 1.58 | 0.23 | 1.16 | 0.14, 9.52 | 0.89 |
| Neurosciences ICU | 0.46 | 0.12, 1.74 | 0.25 | 2.92 | 0.21, 40.53 | 0.42 |
| Surgical ICU | 0.14 | 0.03, 0.71 | 0.02 | 0.04 | 0.00, 1.14 | 0.06 |
| *Primary ICU admission diagnosis* |  |  |  |  |  |  |
| Burn | 0.43 | 0.06, 3.27 | 0.41 | 1.84 | 0.03, 113.30 | 0.77 |
| Cardiovascular | 0.44 | 0.09, 2.18 | 0.31 | 0.42 | 0.03, 5.41 | 0.51 |
| Dermatology | 0.57 | 0.05, 6.90 | 0.66 | 0.50 | 0.01, 22.50 | 0.72 |
| Electrolyte Abnormalities | 0.57 | 0.05, 6.90 | 0.66 | 0.52 | 0.02, 13.60 | 0.69 |
| Endocrine | 0.00 | 0.00, Inf | 0.99 | 0.00 | 0.00, Inf | 0.99 |
| Fever | 2.83 | 0.19, 41.99 | 0.45 | 21.13 | 0.25, 1786.36 | 0.18 |
| Gastrointestinal | 0.59 | 0.10, 3.48 | 0.56 | 0.20 | 0.01, 3.26 | 0.26 |
| Hematologic | 6.80 | 0.95, 48.69 | 0.06 | 2.91 | 0.14, 59.18 | 0.49 |
| Hepatic | 4.25 | 0.45, 40.01 | 0.21 | 2.95 | 0.08, 105.83 | 0.55 |
| Infection | 0.43 | 0.04, 5.11 | 0.50 | 0.52 | 0.02, 13.46 | 0.70 |
| Mental Health | 0.00 | 0.00, Inf | 1.00 | 0.00 | 0.0000, Inf | 1.00 |
| Neoplasm | 1.10 | 0.18, 6.62 | 0.92 | 1.30 | 0.10, 16.64 | 0.84 |
| Neurology | 1.00 | 0.20, 4.98 | 1.00 | 0.78 | 0.06, 9.32 | 0.84 |
| Pneumonia | 1.89 | 0.23, 15.74 | 0.56 | 0.93 | 0.05, 17.58 | 0.96 |
| Pregnancy | 0.00 | 0.00, Inf | 0.99 | 0.00 | 0.00, Inf | 1.00 |
| Pulmonary | 2.13 | 0.44, 10.22 | 0.35 | 1.71 | 0.19, 15.65 | 0.64 |
| Renal | 1.00 | 0.13, 7.94 | 1.00 | 0.81 | 0.03, 19.26 | 0.90 |
| Respiratory | 1.06 | 0.08, 13.52 | 0.96 | 1.42 | 0.03, 58.90 | 0.85 |
| Respiratory failure | 4.25 | 0.73, 24.77 | 0.11 | 1.02 | 0.06, 17.13 | 0.99 |
| Sepsis | 1.47 | 0.26, 8.40 | 0.67 | 0.47 | 0.04, 5.38 | 0.54 |
| Shock | 2.13 | 0.25, 17.93 | 0.49 | 0.40 | 0.02, 9.70 | 0.57 |
| Syncope | 0.00 | 0.00, Inf | 0.99 | 0.00 | 0.00, Inf | 1.00 |
| Toxicology/Ingestion | 0.00 | 0.00, Inf | 0.99 | 0.00 | 0.00, Inf | 0.99 |
| Trauma | 0.75 | 0.11, 4.92 | 0.76 | 0.39 | 0.02, 9.24 | 0.56 |
| Weakness | 2.13 | 0.15, 29.66 | 0.58 | 0.31 | 0.00, 47.55 | 0.65 |
| 24 h after ICU admission |  |  |  |  |  |  |
| Severity of illness, mean (SD) |  |  |  |  |  |  |
| APACHE II Score | 1.18 | 1.14, 1.23 | <0.01 | 1.10 | 0.98, 1.22 | 0.10 |
| SOFA Score | 1.30 | 1.23, 1.39 | <0.01 | 1.36 | 1.10, 1.68 | <0.01 |
| Vital Signs |  |  |  |  |  |  |
| Heart rate | 1.02 | 1.01, 1.03 | <0.01 | 1.02 | 1.00, 1.03 | 0.05 |
| Systolic blood pressure abnormal | 2.61 | 1.62, 4.21 | <0.01 | 0.90 | 0.40, 2.05 | 0.80 |
| Temperature | 0.99 | 0.94, 1.03 | 0.50 | 1.03 | 0.91, 1.17 | 0.61 |
| ARDS Classification |  |  |  |  |  |  |
| Mild | 0.63 | 0.23, 1.75 | 0.38 | 0.55 | 0.13, 2.37 | 0.42 |
| Moderate | 2.24 | 0.99, 5.08 | 0.05 | 2.63 | 0.72, 9.60 | 0.14 |
| Severe | 2.67 | 1.00, 7.13 | 0.05 | 1.15 | 0.24, 5.68 | 0.86 |
| Supportive devices |  |  |  |  |  |  |
| CRRT at 24 h | 3.48 | 0.91, 13.41 | 0.07 | 0.34 | 0.02, 4.68 | 0.42 |
| MV at 24 h | 4.43 | 2.74, 7.17 | <0.01 | 0.77 | 0.18, 3.40 | 0.73 |
| Serum laboratory values |  |  |  |  |  |  |
| Albumin mg/dL | 0.29 | 0.18, 0.46 | <0.01 | 0.56 | 0.25, 1.22 | 0.15 |
| Bicarbonate mEq/L | 0.93 | 0.89, 0.98 | <0.01 | 0.96 | 0.90, 1.04 | 0.32 |
| Creatinine mg/dL | 1.11 | 1.01, 1.23 | 0.04 | 0.89 | 0.69, 1.15 | 0.36 |
| Glucose mg/dL | 1.00 | 1.00, 1.00 | 0.11 | 1.00 | 1.00, 1.00 | 0.81 |
| Lactate mmol/L | 1.19 | 1.06, 1.33 | <0.01 | 0.89 | 0.71, 1.13 | 0.35 |
| Potassium mEq/L | 1.27 | 0.94, 1.71 | 0.12 | 0.74 | 0.48, 1.16 | 0.19 |
| pH < 7.2 | 4.10 | 1.98, 8.50 | <0.01 | 3.02 | 0.77, 11.82 | 0.11 |
| pH > 7.5 | 2.38 | 0.97, 5.80 | 0.06 | 2.14 | 0.54, 8.52 | 0.28 |
| Sodium mEq/L | 0.98 | 0.94, 1.03 | 0.48 | 0.98 | 0.92, 1.04 | 0.45 |
| Hemoglobin g/dL | 0.79 | 0.71, 0.88 | <0.01 | 0.24 | 0.10, 0.60 | <0.01 |
| Hematocrit % | 0.93 | 0.90, 0.97 | <0.01 | 1.51 | 1.11, 2.03 | <0.01 |
| Platelets x 10<sup>3</sup>/μL | 1.00 | 1.00, 1.00 | 0.68 | 1.00 | 1.00, 1.00 | 0.50 |
| White blood cells x 10<sup>3</sup>/μL | 1.07 | 1.04, 1.11 | <0.01 | 1.05 | 0.99, 1.11 | 0.10 |
| Fluid balance at 24 h (mL) | 1.04 | 0.97, 1.12 | 0.30 | 0.96 | 0.78, 1.18 | 0.70 |
| Medications |  |  |  |  |  |  |
| MRC-ICU | 1.08 | 1.05, 1.10 | <0.01 | 0.95 | 0.88, 1.03 | 0.22 |
| Vasopressor at 24 h | 4.02 | 2.50, 6.49 | <0.01 | 0.57 | 0.19, 1.71 | 0.32 |
| Data are presented as n (%) or mean ± standard deviation (SD) unless otherwise stated. |  |  |  |  |  |  |

APACHE: acute physiology and chronic health evaluation; CRRT: continuous renal replacement therapy; ICU: intensive care unit; MRC-ICU: medication regimen complexity-ICU; MV: mechanical ventilation; SOFA: sequential organ failure assessment

To assess both the specific and overall impact of each predictor, we performed two types of regression analyses: 1) individual logistic regressions examining a single variable alongside its relevant missingness indicator (if applicable), and 2) a comprehensive additive logistic regression encompassing all predictors and their respective missingness indicators. During the model training, 5-fold cross validation was applied to choose the hyperparameters for local linear logistic regression and the three machine leaning models.

With the optimal hyperparameters selected, the models were then fitted again on the training set. For local linear logistic regression, the proportion of observations in a neighborhood of every point to be used for fitting the models was tuned. For Random Forest, two hyperparameters were tuned (number of trees and number of variables randomly sampled as candidates at each split). For SVM, linear kernel was used, and cost of constraints violation was tuned. For XGBoost, two hyperparameters were tuned (maximum depth of a tree and maximum number of boosting iterations). Predictions for mortality for all 7 models were made using the test set.

Based on a previous study,[11] the baseline model was a logistic regression model including APACHE II, SOFA, and MRC-ICU as predictors, benchmarked against models including only SOFA or APACHE II as predictor variables. To evaluate the predictive abilities of each model on hospital mortality, area under the receiver operating characteristic curve (AUROC) was computed in addition to sensitivity, specificity, negative predictive value (NPV), positive predictive value (PPV) (or precision), and accuracy in the test set. Results were subsequently compared between AUROCs using DeLong’s test where prediction thresholds were chosen by maximizing Informedness, Matthew’s Correlation Coefficient (MCC) and F1 scores in the training set. A two-sided p-value less than 0.05 was used to determine statistical significance for all variables. All analyses were performed using *R* (version 4.3.0).

### Validation

To verify the models’ performance, we validated them using the UNC 5000 dataset. This involved applying our trained models to the validation data, generating ROC curves, and assessing model metrics.

## Results

Of the initial 1,000 patients randomly selected for inclusion, 9 patients were excluded because the ICU admission from which data was drawn did not represent the index ICU admission. The remaining 991 patients were included in the final cohort (Supplemental Digital Content – Figure 1). The population was 56.81% male and an average of 61.24 years old (SD 17.58). Predominantly represented ICUs included cardiac (30.78%) and medical (40.77%), with cardiovascular (24.62%), neurology (12.21%), and pulmonary (8.88%) representing the most common ICU admission diagnosis categories. Of the total cohort, 9.79% (n = 97) experienced the primary outcome of hospital mortality. Demographic characteristics for the entire cohort as well as cohorts stratified by mortality outcome are described in **Table 1**.

**Figure 1.**
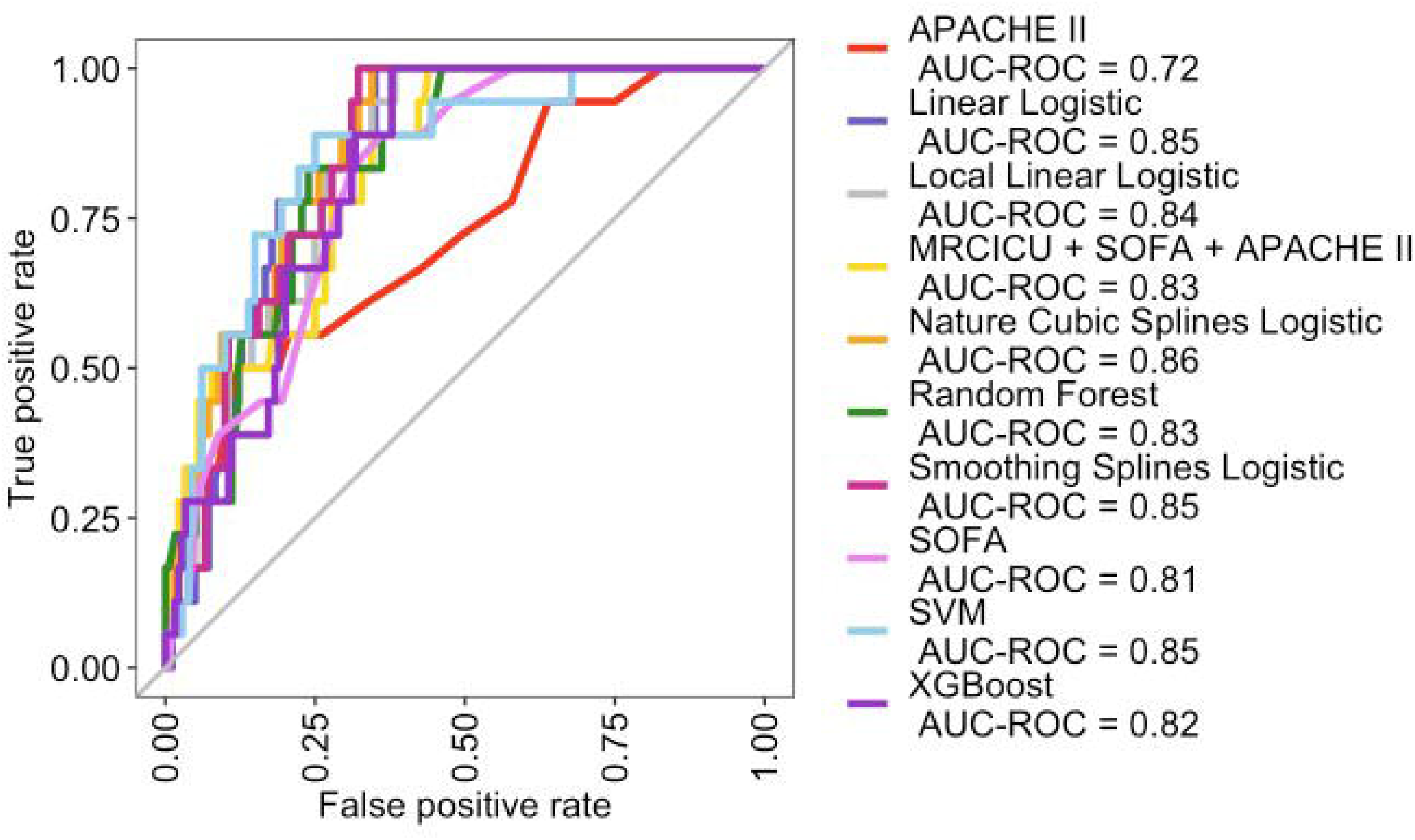
AUROCs for hospital mortality prediction on test set

Univariate and multivariate analysis of each predictor variable is provided in **Table 2**. While several variables had statistically significant associations with mortality on univariate analysis, only sex (as a baseline variable), SOFA score, heart rate, and hemoglobin/hematocrit (as clinical/laboratory variables within the first 24 hours of ICU admission) maintained a statistically significant association in multivariate analysis. After stepwise selection, the four regression models involved similar predictors, which are given in **Table 3** together with the corresponding p values. Notably, lowest SOFA score at 24 hours, lowest hematocrit at 24 hours, and an indicator of missingness for temperature at 24 hours were present in all models, with age and lowest albumin at 24 hours included in three of the models and fluid balance included in two of the models based on ANOVA for nonparametric effects. The local linear logistic regression after stepwise selection did not have higher order terms and used the same predictors as the logistic regression with linear predictors.

**Table 3.** Regression for mortality with linear predictors.

| Linear predictors |  |  |  |  |
| --- | --- | --- | --- | --- |
|  | OR | 95% CI | p-value |  |
| SOFA at 24 hours | 1.28 | 1.19, 1.37 | <0.01 |  |
| Age | 1.04 | 1.02, 1.06 | <0.01 |  |
| Indicator for missingness of temperature at 24 hours | 5.21 | 1.62, 17.00 | <0.01 |  |
| Lowest albumin at 24 hours | 0.41 | 0.28, 0.59 | <0.01 |  |
| Lowest HGB at 24 hours | 0.41 | 0.24, 0.69 | <0.01 |  |
| Lowest HCT at 24 hours | 1.36 | 1.14, 1.62 | <0.01 |  |
| Nature cubic splines |  |  |  |  |
|  | OR | 95% CI | p-value |  |
| SOFA at 24 hours (Linear predictor) | 1.24 | 1.11, 1.38 | <0.01 |  |
| APACHE II at 24 hours (Nature cubic spline with knots at 8.5 and 16.5) |  |  |  |  |
| Basis 1 | 7.23*10<sup>2</sup> | 11.26, 3.86*10<sup>5</sup> | 0.04 |  |
| Basis 2 | 5.61*10<sup>5</sup> | 0.51, 8.77*10<sup>17</sup> | 0.21 |  |
| Basis 3 | 3.58*10<sup>2</sup> | 6.26, 5.10*10<sup>5</sup> | 0.03 |  |
| Mechanical Ventilation at 24 hours | 0.25 | 0.10, 0.63 | <0.01 |  |
| Indicator for missingness of temperature at 24 hours | 5.94 | 1.81, 20.04 | <0.01 |  |
| Indicator for missingness of albumin at 24 hours | 0.32 | 0.17, 0.60 | <0.01 |  |
| Lowest HGB at 24 hours (Nature cubic spline with knots at 12.5 and 14.1) |  |  |  |  |
| Basis 1 | 0.00 | 0.00, 1.44*10<sup>-3</sup> | <0.01 |  |
| Basis 2 | 0.00 | 0.00, 9.33*10<sup>-4</sup> | <0.01 |  |
| Basis 3 | 0.43 | 0.00, 1.43*10<sup>4</sup> | 0.86 |  |
| Lowest HCT at 24 hours (Nature cubic spline with knots at 40.6 and 42.4) |  |  |  |  |
| Basis 1 | 2.50*10<sup>4</sup> | 84.54, 7.53*10<sup>6</sup> | <0.01 |  |
| Basis 2 | 1.86*10<sup>5</sup> | 0.00, 1.48*10<sup>13</sup> | 0.23 |  |
| Basis 3 | 0.00 | 0.00, 6.67*10<sup>2</sup> | 0.24 |  |
| Smoothing splines |  |  |  |  |
| ANOVA for parametric effects | Df | MSE | F-value | p-value |
| SOFA at 24 hours | 1 | 52.65 | 73.74 | <0.01 |
| Age | 1 | 4.86 | 6.80 | <0.01 |
| Fluid balance at 24 hours (Smoothing spline with df=2) | 1 | 0.67 | 0.93 | 0.33 |
| Indicator for missingness of temperature at 24 hours | 1 | 8.40 | 11.77 | <0.01 |
| Lowest albumin at 24 hours | 1 | 29.39 | 41.17 | <0.01 |
| Lowest HGB at 24 hours | 1 | 0.01 | 0.01 | 0.91 |
| Lowest HCT at 24 hours | 1 | 11.35 | 15.89 | <0.01 |
| Residuals | 784 | 0.714 |  |  |
| ANOVA for nonparametric effects | Df |  | Chisq-Value | p-value |
| Fluid balance at 24 hours (Smoothing spline with df=2) | 1 |  | 7.18 | <0.01 |
| Local linear |  |  |  |  |
| ANOVA for parametric effects | Df | MSE | F-value | p-value |
| SOFA at 24 hours | 1 | 53.99 | 74.54 | <0.01 |
| Age | 1 | 4.93 | 6.81 | <0.01 |
| Fluid balance at 24 hours (local linear predictor) | 1 | 0.58 | 0.81 | 0.37 |
| Indicator for missingness of temperature at 24 hours | 1 | 9.01 | 12.44 | <0.01 |
| Lowest albumin at 24 hours | 1 | 29.94 | 41.34 | <0.01 |
| Lowest HGB at 24 hours | 1 | 0.02 | 0.02 | 0.88 |
| Lowest HCT at 24 hours | 1 | 11.43 | 15.77 | <0.01 |
| Residuals | 783.69 | 0.724 |  |  |
| **ANOVA for nonparametric effects** | **Df** | **Chisq-Value** | **p-value** |  |
| Fluid balance at 24 hours (Smoothing spline with df=2) | 1.3 | 10.31 | <0.01 |  |
| APACHE: acute physiology and chronic health evaluation; HCT: hematocrit; HGB: hemoglobin; SOFA: sequential organ failure assessment |  |  |  |  |

AUROC curves were plotted for ten models (**Figure 1**). Adding medication data (MRC-ICU) to a model including SOFA and APACHE II as clinical severity of illness metrics and mortality predictors increased the AUROC compared to benchmarking models based on APACHE II and SOFA alone (0.83 vs 0.72 and 0.81, respectively). A model inclusive of SOFA, APACHE II, Mechanical Ventilation, Albumin, Temperature, Hemoglobin, and Hematocrit increased the AUROC to 0.86. AUROC and their corresponding confidence intervals for each model are provided in **Table 4**. Detailed model performance metrics including accuracy, sensitivity, specificity, PPV, and NPV values on the test set with thresholds chosen by different criteria are reported in **Table 5**.

**Table 4.** AUROC for mortality prediction models on test set.

|  | **AUROC** |
| --- | --- |
| *APACHE II* | 0.72, 0.58-0.86 |
| *SOFA* | 0.81, 0.69-0.93 |
| *MRC-ICU + SOFA + APACHE II* | 0.83, 0.71-0.95 |
| *Linear Logistic* | 0.85, 0.74-0.96 |
| *Nature Cubic Splines Logistic* | 0.86, 0.75-0.97 |
| *Smoothing Splines Logistic* | 0.85, 0.74-0.96 |
| *Local Linear Logistic* | 0.84, 0.72-0.96 |
| *Random Forest* | 0.83, 0.71-0.95 |
| *SVM* | 0.85, 0.74-0.96 |
| *XGBoost* | 0.82, 0.7-0.94 |
| APACHE: acute physiology and chronic health evaluation; AUROC: area under the receiver operating characteristic; SOFA: sequential organ failure assessment |  |

**Table 5.**
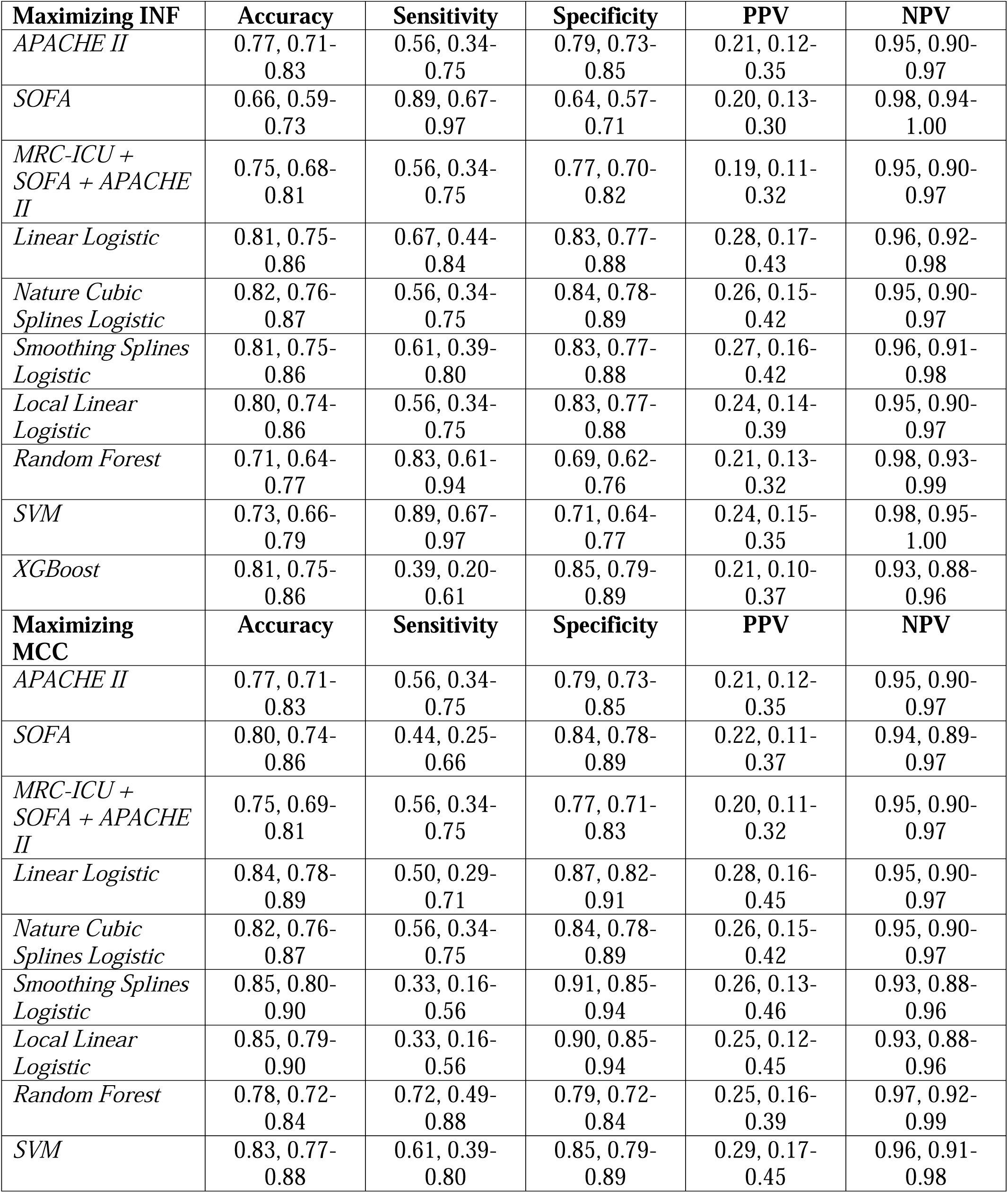
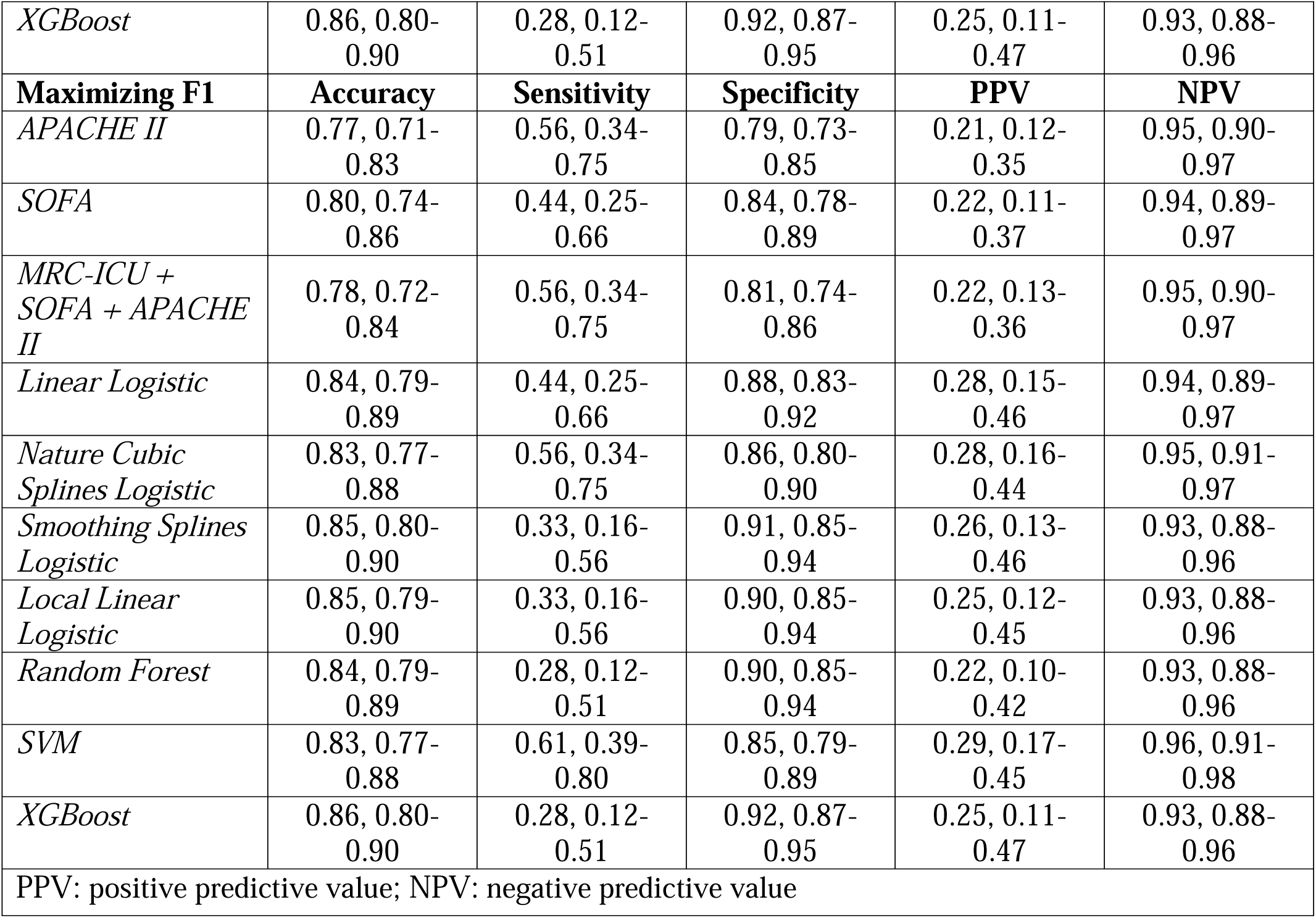
Accuracy, sensitivity, specificity, negative predictive value, and positive predictive value for mortality prediction models on test set.

| **Maximizing INF** | **Accuracy** | **Sensitivity** | **Specificity** | **PPV** | **NPV** |
| --- | --- | --- | --- | --- | --- |
| *APACHE II* | 0.77, 0.71-0.83 | 0.56, 0.34-0.75 | 0.79, 0.73-0.85 | 0.21, 0.12-0.35 | 0.95, 0.90-0.97 |
| *SOFA* | 0.66, 0.59-0.73 | 0.89, 0.67-0.97 | 0.64, 0.57-0.71 | 0.20, 0.13-0.30 | 0.98, 0.94-1.00 |
| *MRC-ICU + SOFA + APACHE II* | 0.75, 0.68-0.81 | 0.56, 0.34-0.75 | 0.77, 0.70-0.82 | 0.19, 0.11-0.32 | 0.95, 0.90-0.97 |
| *Linear Logistic* | 0.81, 0.75-0.86 | 0.67, 0.44-0.84 | 0.83, 0.77-0.88 | 0.28, 0.17-0.43 | 0.96, 0.92-0.98 |
| *Nature Cubic Splines Logistic* | 0.82, 0.76-0.87 | 0.56, 0.34-0.75 | 0.84, 0.78-0.89 | 0.26, 0.15-0.42 | 0.95, 0.90-0.97 |
| *Smoothing Splines Logistic* | 0.81, 0.75-0.86 | 0.61, 0.39-0.80 | 0.83, 0.77-0.88 | 0.27, 0.16-0.42 | 0.96, 0.91-0.98 |
| *Local Linear Logistic* | 0.80, 0.74-0.86 | 0.56, 0.34-0.75 | 0.83, 0.77-0.88 | 0.24, 0.14-0.39 | 0.95, 0.90-0.97 |
| *Random Forest* | 0.71, 0.64-0.77 | 0.83, 0.61-0.94 | 0.69, 0.62-0.76 | 0.21, 0.13-0.32 | 0.98, 0.93-0.99 |
| *SVM* | 0.73, 0.66-0.79 | 0.89, 0.67-0.97 | 0.71, 0.64-0.77 | 0.24, 0.15-0.35 | 0.98, 0.95-1.00 |
| *XGBoost* | 0.81, 0.75-0.86 | 0.39, 0.20-0.61 | 0.85, 0.79-0.89 | 0.21, 0.10-0.37 | 0.93, 0.88-0.96 |
| **Maximizing MCC** | **Accuracy** | **Sensitivity** | **Specificity** | **PPV** | **NPV** |
| *APACHE II* | 0.77, 0.71-0.83 | 0.56, 0.34-0.75 | 0.79, 0.73-0.85 | 0.21, 0.12-0.35 | 0.95, 0.90-0.97 |
| *SOFA* | 0.80, 0.74-0.86 | 0.44, 0.25-0.66 | 0.84, 0.78-0.89 | 0.22, 0.11-0.37 | 0.94, 0.89-0.97 |
| *MRC-ICU + SOFA + APACHE II* | 0.75, 0.69-0.81 | 0.56, 0.34-0.75 | 0.77, 0.71-0.83 | 0.20, 0.11-0.32 | 0.95, 0.90-0.97 |
| *Linear Logistic* | 0.84, 0.78-0.89 | 0.50, 0.29-0.71 | 0.87, 0.82-0.91 | 0.28, 0.16-0.45 | 0.95, 0.90-0.97 |
| *Nature Cubic Splines Logistic* | 0.82, 0.76-0.87 | 0.56, 0.34-0.75 | 0.84, 0.78-0.89 | 0.26, 0.15-0.42 | 0.95, 0.90-0.97 |
| *Smoothing Splines Logistic* | 0.85, 0.80-0.90 | 0.33, 0.16-0.56 | 0.91, 0.85-0.94 | 0.26, 0.13-0.46 | 0.93, 0.88-0.96 |
| *Local Linear Logistic* | 0.85, 0.79-0.90 | 0.33, 0.16-0.56 | 0.90, 0.85-0.94 | 0.25, 0.12-0.45 | 0.93, 0.88-0.96 |
| *Random Forest* | 0.78, 0.72-0.84 | 0.72, 0.49-0.88 | 0.79, 0.72-0.84 | 0.25, 0.16-0.39 | 0.97, 0.92-0.99 |
| *SVM* | 0.83, 0.77-0.88 | 0.61, 0.39-0.80 | 0.85, 0.79-0.89 | 0.29, 0.17-0.45 | 0.96, 0.91-0.98 |
| *XGBoost* | 0.86, 0.80-0.90 | 0.28, 0.12-0.51 | 0.92, 0.87-0.95 | 0.25, 0.11-0.47 | 0.93, 0.88-0.96 |
| **Maximizing F1** | **Accuracy** | **Sensitivity** | **Specificity** | **PPV** | **NPV** |
| *APACHE II* | 0.77, 0.71-0.83 | 0.56, 0.34-0.75 | 0.79, 0.73-0.85 | 0.21, 0.12-0.35 | 0.95, 0.90-0.97 |
| *SOFA* | 0.80, 0.74-0.86 | 0.44, 0.25-0.66 | 0.84, 0.78-0.89 | 0.22, 0.11-0.37 | 0.94, 0.89-0.97 |
| *MRC-ICU + SOFA + APACHE II* | 0.78, 0.72-0.84 | 0.56, 0.34-0.75 | 0.81, 0.74-0.86 | 0.22, 0.13-0.36 | 0.95, 0.90-0.97 |
| *Linear Logistic* | 0.84, 0.79-0.89 | 0.44, 0.25-0.66 | 0.88, 0.83-0.92 | 0.28, 0.15-0.46 | 0.94, 0.89-0.97 |
| *Nature Cubic Splines Logistic* | 0.83, 0.77-0.88 | 0.56, 0.34-0.75 | 0.86, 0.80-0.90 | 0.28, 0.16-0.44 | 0.95, 0.91-0.97 |
| *Smoothing Splines Logistic* | 0.85, 0.80-0.90 | 0.33, 0.16-0.56 | 0.91, 0.85-0.94 | 0.26, 0.13-0.46 | 0.93, 0.88-0.96 |
| *Local Linear Logistic* | 0.85, 0.79-0.90 | 0.33, 0.16-0.56 | 0.90, 0.85-0.94 | 0.25, 0.12-0.45 | 0.93, 0.88-0.96 |
| *Random Forest* | 0.84, 0.79-0.89 | 0.28, 0.12-0.51 | 0.90, 0.85-0.94 | 0.22, 0.10-0.42 | 0.93, 0.88-0.96 |
| *SVM* | 0.83, 0.77-0.88 | 0.61, 0.39-0.80 | 0.85, 0.79-0.89 | 0.29, 0.17-0.45 | 0.96, 0.91-0.98 |
| *XGBoost* | 0.86, 0.80-0.90 | 0.28, 0.12-0.51 | 0.92, 0.87-0.95 | 0.25, 0.11-0.47 | 0.93, 0.88-0.96 |
| PPV: positive predictive value; NPV: negative predictive value |  |  |  |  |  |

Machine learning models (Random Forest, SVM, XGBoost) provided similar AUROC for hospital mortality prediction (0.83, 0.85, and 0.82, respectively) as traditional regression methods as well as the model based on MRC-ICU + SOFA + APACHE II. Feature importance graphs for XGBoost and Random Forest models reveal similar contributing variables (**Figure 2** and Supplemental Digital Content – Figure 2), with SOFA, APACHE II, Age, Heart Rate, and some laboratory values prominently featured.

**Figure 2.**
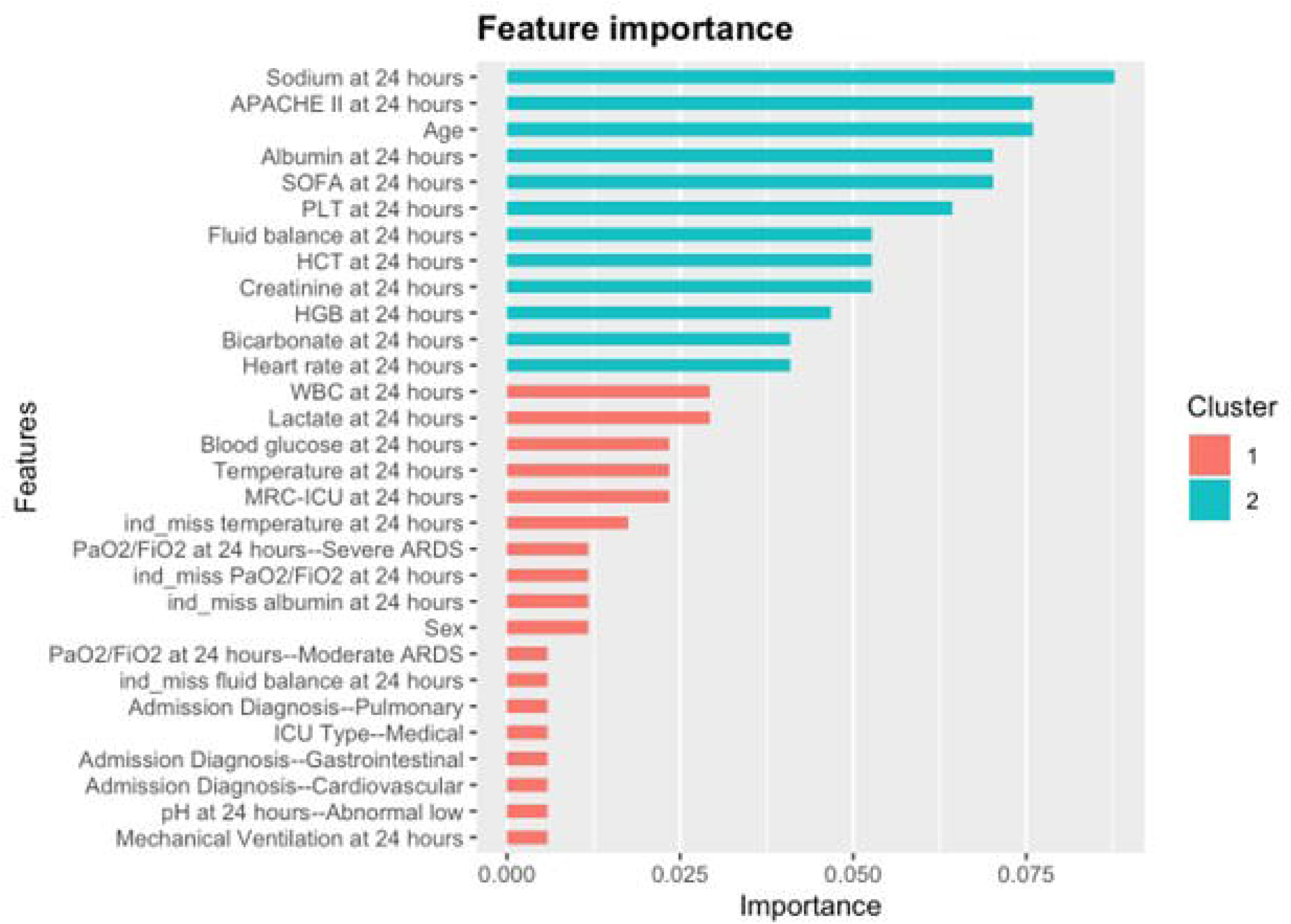
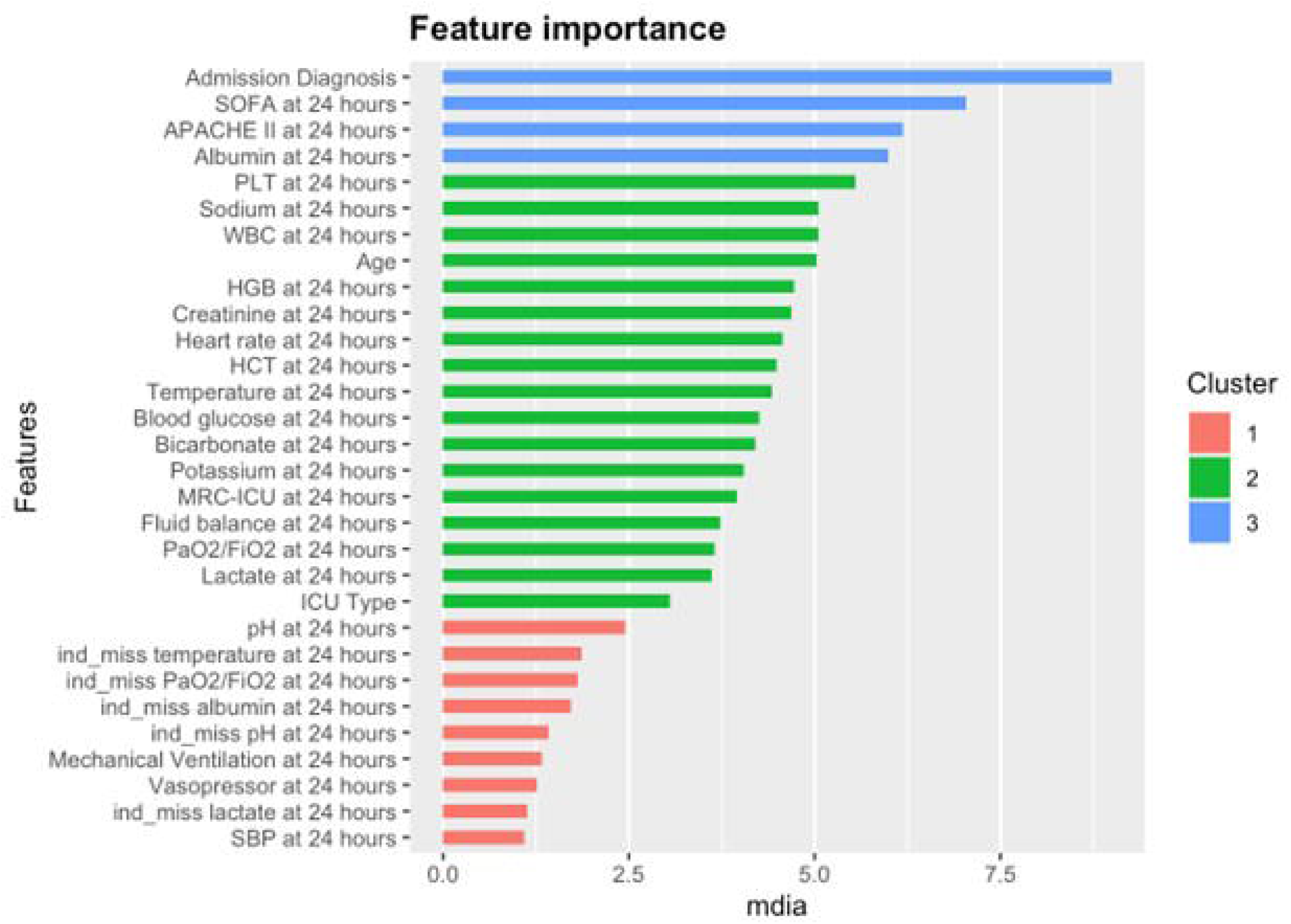
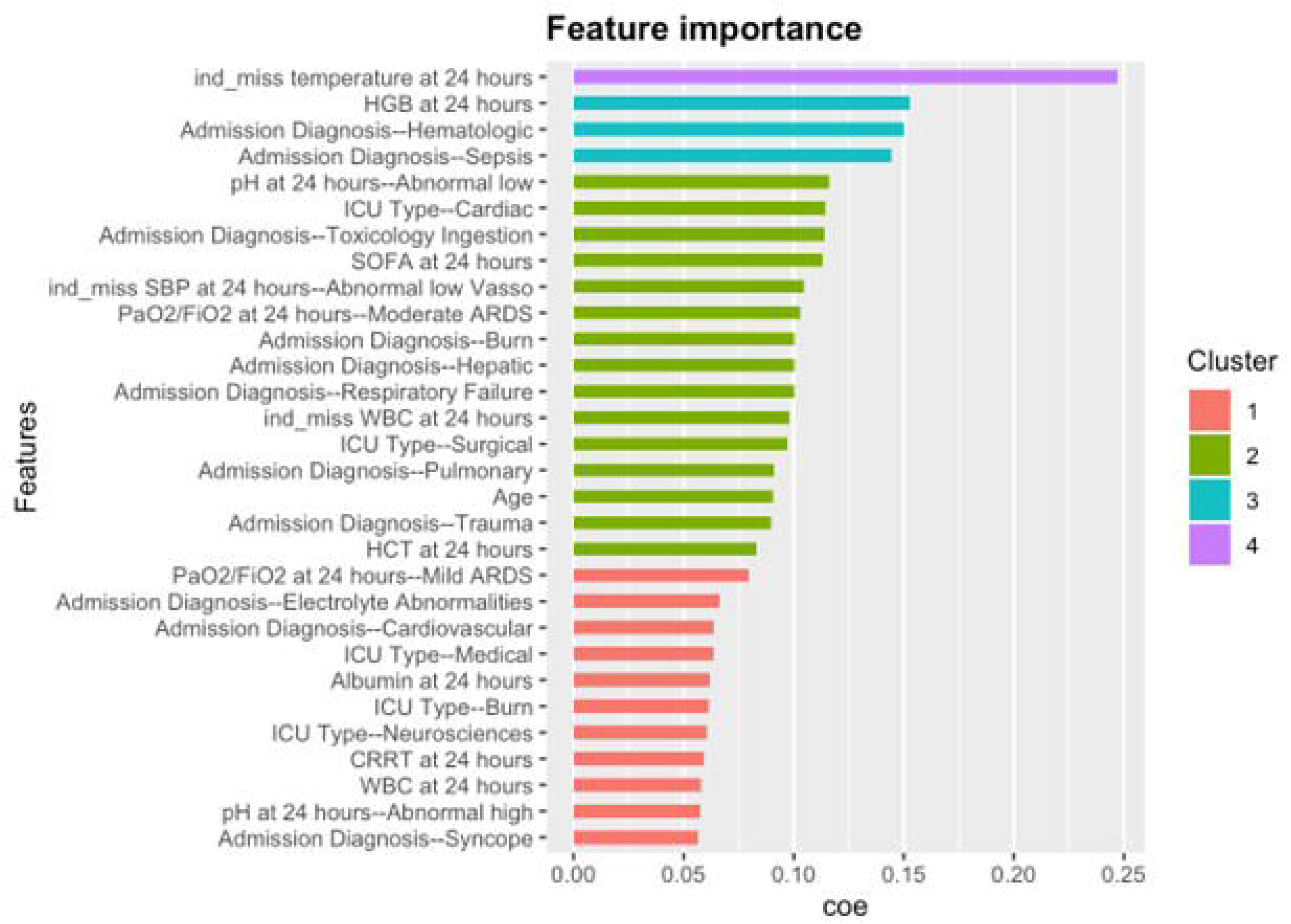
Feature importance for mortality prediction with XGBoost

Medication data in the form of MRC-ICU demonstrated feature importance similar to serum lactate or ARDS classification and ranked higher than presence of mechanical ventilation in the first 24 hours. Feature importance graphs generated for SVM models did not feature MRC-ICU (Supplemental Digital Content – Figure 3).

Validation was performed on a temporally separate dataset which, after excluding patients for whom data was not from the index ICU admission, included a total of 4,878 patients. The population was 51.91% male with an average age of 58.87 years old (SD 16.47). Compared to the cohort from which the training and test sets were derived, the validation cohort featured a greater percentage of medical ICU admissions (94.94%) and had fewer admissions related to a primary cardiovascular diagnosis (7.46%), with a greater proportion of respiratory or respiratory failure (16.73%), sepsis (11.46%), gastrointestinal (11.75%), and infection-related admissions (8.18%). Of the total validation cohort, 19.76% (n = 964) experienced the primary outcome of hospital mortality. Demographic characteristics for the entire validation cohort as well as cohorts stratified by mortality outcome are described in Supplemental Digital Content – Table 3. Missingness characteristics for each of the variables in the validation cohort is provided in Supplemental Digital Content – Table 4.

Each of the trained models was applied to the validation cohort, with AUROC curves generated (Supplemental Digital Content – Figure 4). Model performance on the validation cohort was consistently lower than performance on the test set across all models (Supplemental Digital Content – Tables 5 and 6). Traditional regression methods and models built on severity of illness metrics and medication-related data performed as well as machine learning-based models in the validation cohort. The Random Forest model exhibited the most stable performance from the test set to the validation set.

## Discussion

The prediction of ICU outcomes including mortality remains an ongoing area of research, with new models and scores developed frequently.[27–29] However, model performance varies and even commonly used mortality predictors suffer from shortcomings.[30] Proposed methods for model optimization include customization of scores using local data[31] and the use of AI and machine learning methods to identify complex interactions and associations not apparent in traditional modeling.[14, 29, 32–35] It is notable that contemporary mortality prediction scores and metrics (e.g., SOFA, APACHE) do not include medication data (beyond vasoactive agents), despite the fact that medications are known to be causal agents both for positive outcomes via the treatment of disease as well as negative outcomes via both predictable and unpredictable interactions with the patient, the disease, or the medication regimen.[1, 3, 5]

Addition of medication data (as incorporated into the MRC-ICU score, a summary characterization of medication use in the ICU) to severity of illness scores (i.e., SOFA, APACHE II) in traditional modeling improved performance for hospital mortality prediction in this study; however, MRC-ICU was not identified as a significant predictor in logistic regression models. MRC-ICU has previously been shown to be associated with mortality,[36–38] number and intensity of required medication interventions,[37, 39] need for mechanical ventilation,[10] and development of fluid overload.[40] Addition of MRC-ICU to conventional severity of illness scores (i.e., SOFA, APACHE) with traditional modeling techniques improved mortality prediction,[11] although there was a positive association between MRC-ICU and mortality in univariate analysis and a negative association in multivariate analysis inclusive of severity of illness metrics, suggesting a complex modifying relationship.

Machine learning-based models did not out-perform traditional modeling methods using simple and easily retrievable variables, but machine learning approaches may be useful for further elucidating complex relationships between variables or causal relationships behind associations in traditional modeling. Interestingly, MRC-ICU had similar feature importance to clinical variables such as lactate and fluid balance in the prediction of hospital mortality. Machine learning approaches may be particularly useful for the incorporation of medication data, which is problematic for traditional modeling because of the large number of variables involved. Inclusion of MRC-ICU in machine learning models has shown the ability to improve prediction of patient outcomes compared to traditional modeling methods, albeit modestly.[10, 41]

An interesting finding from this study is that machine learning methods identified different predictors of mortality than traditional logistic regression models (and even than models based on different machine learning models). This finding highlights the potential value of machine learning approaches for identifying and characterizing previously unknown associations, but also emphasizes the “black box” nature of such an approach. Despite the potential impact of AI and machine learning approaches, it is important to consider whether performance of these models significantly improves upon traditional models that are simpler and more transparent, considering the resources required to implement these approaches.[9, 42] This study also highlights the need for prediction models to benchmark against existing clinical standards.

When models generated on a training set of nearly 1,000 patients were validated against a set of nearly 5,000 patients, AUROCs were consistently lower than they were on the test set. This discrepancy in performance may stem from structural differences between the two datasets, which could be initially discernible through their characteristics. Determining which dataset better represents the population is challenging. However, the current model results underscore the utility of the models in predicting mortality as well as the need for institution or population specific evaluation of a base model.

This study has notable limitations. While sample sizes of nearly 1,000 and 5,000 patients are large for most critical care studies, these sample sizes are small for the purposes of machine learning. Small sample sizes may lead to model overfitting and a lack of generalizability. While handling of missingness in this study was consistent, the use of normal values to substitute for missing data may have altered model results. Bias may exist as a result of which patients had specific data points available. It is possible that data points not collected could improve model performance for hospital mortality prediction. Limiting model metrics to the admission timepoint and the first 24 hours may not capture all potential contributors to and predictors of hospital mortality and limits assessment of the dynamic nature of critical illness over the course of an ICU stay. Hospital mortality as an outcome also has potential limitations, and using a specific timepoint may have altered outcomes of this study. Finally, all data sets in this study include patients from a single health system and may not be readily generalizable to patients in other health systems or geographic regions.

This study represents the largest and most robust evaluation to date of the MRC-ICU score with a training/test set of nearly 1,000 patients and a validation set of nearly 5,000 patients. It is the largest study to date incorporating medication regimen complexity, severity of illness data, and machine learning methods evaluating a patient-centered outcome that is relevant to potential future applications. Given these results, future studies will focus on the use of larger datasets with more granular medication data, as these features represent modifiable components in a causal model of patient outcomes.

## Conclusion

Accurate prediction of mortality is an ongoing challenge in critically ill patient populations. Medications are known to be causal agents for mortality outcomes. Incorporation of medication data has potential promise to improve the performance of mortality prediction models in the ICU. While machine learning approaches may improve prediction performance in some settings, performance should be benchmarked against simpler and more transparent models for outcome prediction.

## Supporting information

Supplemental Content

## Data Availability

All data produced in the present study are available upon reasonable request to the authors

## Acknowledgements

Data acquisition were supported by NC Tracs, funded by Grant Number UL1TR002489 from the National Center for Advancing Translations Sciences at the National Institutes of Health, and Data Analytics at the University of North Carolina Medical Center Department of Pharmacy.

## Declarations Ethical Approval

This was an observational study that was reviewed by the University of Georgia Institutional Review Board (IRB) and determined to be exempt from IRB oversight (Project00001541).

## Author Contributions

BM conducted manuscript drafting, results interpretation, and critical revisions. TZ and XC conducted data pre-processing, data analysis, and methods development. AM, SS, JD, RK, and DM provided manuscript review and revisions and clinical results interpretation. AS provided project oversight, manuscript drafting, revisions, and results interpretation.

## Availability of data & materials

Following request and authorship team approval of the appropriateness of the request, datasets can be made available. Please contact the corresponding author.

## Notes

**Conflicts of Interest:** The authors have no conflicts of interest.

### Competing Interest Statement

The authors have declared no competing interest.

### Funding Statement

Funding through Agency of Healthcare Research and Quality for Drs. Devlin, Murphy, Sikora, Smith, and Kamaleswaran was provided through R21HS028485 and R01HS029009.

### Author Declarations

University of Georgia Institutional Review Board

## References

1. Sikora A: Critical Care Pharmacists: A Focus on Horizons. Crit Care Clin 2023, 39(3):503–527.

2. Cullen DJ, Sweitzer BJ, Bates DW, Burdick E, Edmondson A, Leape LL: Preventable adverse drug events in hospitalized patients: a comparative study of intensive care and general care units. Crit Care Med 1997, 25(8):1289–1297.

3. Kane-Gill SL, Kirisci L, Verrico MM, Rothschild JM: Analysis of risk factors for adverse drug events in critically ill patients*. Crit Care Med 2012, 40(3):823–828.

4. Fitzmaurice MG, Wong A, Akerberg H, Avramovska S, Smithburger PL, Buckley MS, Kane-Gill SL: Evaluation of Potential Drug-Drug Interactions in Adults in the Intensive Care Unit: A Systematic Review and Meta-Analysis. Drug Saf 2019, 42(9):1035–1044.

5. Bakker T, Abu-Hanna A, Dongelmans DA, Vermeijden WJ, Bosman RJ, de Lange DW, Klopotowska JE, de Keizer NF, Hendriks S, Ten Cate J et al: Clinically relevant potential drug-drug interactions in intensive care patients: A large retrospective observational multicenter study. J Crit Care 2021, 62:124–130.

6. Giraud T, Dhainaut JF, Vaxelaire JF, Joseph T, Journois D, Bleichner G, Sollet JP, Chevret S, Monsallier JF: Iatrogenic complications in adult intensive care units: a prospective two-center study. Crit Care Med 1993, 21(1):40–51.

7. Rothschild JM, Landrigan CP, Cronin JW, Kaushal R, Lockley SW, Burdick E, Stone PH, Lilly CM, Katz JT, Czeisler CA et al: The Critical Care Safety Study: The incidence and nature of adverse events and serious medical errors in intensive care. Crit Care Med 2005, 33(8):1694–1700.

8. Classen DC, Pestotnik SL, Evans RS, Lloyd JF, Burke JP: Adverse drug events in hospitalized patients. Excess length of stay, extra costs, and attributable mortality. Jama 1997, 277(4):301–306.

9. Sikora A, Zhang T, Murphy DJ, Smith SE, Murray B, Kamaleswaran R, Chen X, Buckley MS, Rowe S, Devlin JW: Machine learning vs. traditional regression analysis for fluid overload prediction in the ICU. Sci Rep 2023, 13(1):19654.

10. Sikora A, Zhao B, Kong Y, Murray B, Shen Y: Machine learning based prediction of prolonged duration of mechanical ventilation incorporating medication data. medRxiv 2023.

11. Sikora A, Devlin JW, Yu M, Zhang T, Chen X, Smith SE, Murray B, Buckley MS, Rowe S, Murphy DJ: Evaluation of medication regimen complexity as a predictor for mortality. Sci Rep 2023, 13(1):10784.

12. Wheelwright J, Halstead ES, Knehans A, Bonavia AS: Ex Vivo Endotoxin Stimulation of Blood for Predicting Survival in Patients With Sepsis: A Systematic Review. CHEST Crit Care 2023, 1(3).

13. Rilinger J, Book R, Kaier K, Giani M, Fumagalli B, Jäckel M, Bemtgen X, Zotzmann V, Biever PM, Foti G et al: A Mortality Prediction Score for Patients With Veno-Venous Extracorporeal Membrane Oxygenation (VV-ECMO): The PREDICT VV-ECMO Score. Asaio j 2023.

14. Yamga E, Mantena S, Rosen D, Bucholz EM, Yeh RW, Celi LA, Ustun B, Butala NM: Optimized Risk Score to Predict Mortality in Patients With Cardiogenic Shock in the Cardiac Intensive Care Unit. J Am Heart Assoc 2023, 12(13):e029232.

15. Keegan MT, Gajic O, Afessa B: Severity of illness scoring systems in the intensive care unit. Crit Care Med 2011, 39(1):163–169.

16. Kramer AA, Zimmerman JE, Knaus WA: Severity of Illness and Predictive Models in Society of Critical Care Medicine’s First 50 Years: A Tale of Concord and Conflict. Crit Care Med 2021, 49(5):728–740.

17. Rafiei A, Rad MG, Sikora A, Kamaleswaran R: Improving irregular temporal modeling by integrating synthetic data to the electronic medical record using conditional GANs: a case study of fluid overload prediction in the intensive care unit. medRxiv 2023.

18. von Elm E, Altman DG, Egger M, Pocock SJ, Gøtzsche PC, Vandenbroucke JP: The Strengthening the Reporting of Observational Studies in Epidemiology (STROBE) statement: guidelines for reporting observational studies. Ann Intern Med 2007, 147(8):573–577.

19. Liu X, Cruz Rivera S, Moher D, Calvert MJ, Denniston AK: Reporting guidelines for clinical trial reports for interventions involving artificial intelligence: the CONSORT-AI extension. Nat Med 2020, 26(9):1364–1374.

20. Schwarz G: Estimating the dimension of a model. The Annals of Statistics 1978, 6(2):461–464.

21. Chen T, Guestrin C: XGBoost: A Scalable Tree Boosting System. In: Proceedings of the 22nd ACM SIGKDD International Conference on Knowledge Discovery and Data Mining. San Francisco, California, USA: Association for Computing Machinery; 2016: 785–794.

22. Cortes C, Vapnik V: Support-vector networks. Machine Learning 1995, 20(3):273–297.

23. Ho TK: Random decision forests. In: Proceedings of the Third International Conference on Document Analysis and Recognition (Volume 1) - Volume 1. IEEE Computer Society; 1995: 278.

24. Liaw A, Wiener MC: Classification and Regression by randomForest. In: 2007; 2007.

25. _e1071: Misc Functions of the Department of Statistics, Probability Theory Group (Formerly: E1071), TU Wien_. R package version 1.7-13, <https://CRAN.R-project.org/package=e1071>

26. _ xgboost: Extreme Gradient Boosting. R package version 1.7.5.1, <https://CRAN.R-project.org/package=xgboost>

27. Keuning BE, Kaufmann T, Wiersema R, Granholm A, Pettilä V, Møller MH, Christiansen CF, Castela Forte J, Snieder H, Keus F et al: Mortality prediction models in the adult critically ill: A scoping review. Acta Anaesthesiol Scand 2020, 64(4):424–442.

28. Rilinger J, Book R, Kaier K, Giani M, Fumagalli B, Jäckel M, Bemtgen X, Zotzmann V, Biever PM, Foti G et al: A Mortality Prediction Score for Patients With Veno-Venous Extracorporeal Membrane Oxygenation (VV-ECMO): The PREDICT VV-ECMO Score. Asaio j 2023, 70(4):293–298.

29. Peres Bota D, Melot C, Lopes Ferreira F, Nguyen Ba V, Vincent JL: The Multiple Organ Dysfunction Score (MODS) versus the Sequential Organ Failure Assessment (SOFA) score in outcome prediction. Intensive Care Med 2002, 28(11):1619–1624.

30. Nassar AP, Jr., Mocelin AO, Nunes AL, Giannini FP, Brauer L, Andrade FM, Dias CA: Caution when using prognostic models: a prospective comparison of 3 recent prognostic models. J Crit Care 2012, 27(4):423.e421–427.

31. Lee J, Maslove DM: Customization of a Severity of Illness Score Using Local Electronic Medical Record Data. J Intensive Care Med 2017, 32(1):38–47.

32. Pirracchio R, Petersen ML, Carone M, Rigon MR, Chevret S, van der Laan MJ: Mortality prediction in intensive care units with the Super ICU Learner Algorithm (SICULA): a population-based study. Lancet Respir Med 2015, 3(1):42–52.

33. Villar J, González-Martín JM, Hernández-González J, Armengol MA, Fernández C, Martín-Rodríguez C, Mosteiro F, Martínez D, Sánchez-Ballesteros J, Ferrando C et al: Predicting ICU Mortality in Acute Respiratory Distress Syndrome Patients Using Machine Learning: The Predicting Outcome and STratifiCation of severity in ARDS (POSTCARDS) Study. Crit Care Med 2023, 51(12):1638–1649.

34. Johnson AE, Kramer AA, Clifford GD: A new severity of illness scale using a subset of Acute Physiology And Chronic Health Evaluation data elements shows comparable predictive accuracy. Crit Care Med 2013, 41(7):1711–1718.

35. Lim L, Gim U, Cho K, Yoo D, Ryu HG, Lee HC: Real-time machine learning model to predict short-term mortality in critically ill patients: development and international validation. Crit Care 2024, 28(1):76.

36. Gwynn ME, Poisson MO, Waller JL, Newsome AS: Development and validation of a medication regimen complexity scoring tool for critically ill patients. Am J Health Syst Pharm 2019, 76(Supplement_2):S34–S40.

37. Sikora A, Ayyala D, Rech MA, Blackwell SB, Campbell J, Caylor MM, Condeni MS, DePriest A, Dzierba AL, Flannery AH et al: Impact of Pharmacists to Improve Patient Care in the Critically Ill: A Large Multicenter Analysis Using Meaningful Metrics With the Medication Regimen Complexity-ICU (MRC-ICU) Score. Crit Care Med 2022, 50(9):1318–1328.

38. Al-Mamun MA, Strock J, Sharker Y, Shawwa K, Schmidt R, Slain D, Sakhuja A, Brothers TN: Evaluating the Medication Regimen Complexity Score as a Predictor of Clinical Outcomes in the Critically Ill. J Clin Med 2022, 11(16).

39. Smith SE, Shelley R, Sikora A: Medication regimen complexity vs patient acuity for predicting critical care pharmacist interventions. Am J Health Syst Pharm 2022, 79(8):651–655.

40. Olney WJ, Chase AM, Hannah SA, Smith SE, Newsome AS: Medication Regimen Complexity Score as an Indicator of Fluid Balance in Critically Ill Patients. J Pharm Pract 2022, 35(4):573–579.

41. Al-Mamun MA, Brothers T, Newsome AS: Development of Machine Learning Models to Validate a Medication Regimen Complexity Scoring Tool for Critically Ill Patients. Ann Pharmacother 2021, 55(4):421–429.

42. Christodoulou E, Ma J, Collins GS, Steyerberg EW, Verbakel JY, Van Calster B: A systematic review shows no performance benefit of machine learning over logistic regression for clinical prediction models. J Clin Epidemiol 2019, 110:12–22.

