## Supplemental Content for "Augmenting mortality prediction with medication data and machine learning models"

**Supplemental Table 1.** Imputed values for features with missingness

| **Feature** | **Feature Type** | **Imputed** |
| --- | --- | --- |
| pH at 24 hours | Categorical | Normal |
| Systolic blood pressure | Categorical | Normal if no Vasopressor used at 24 hours, Abnormal if Vasopressor used at 24 hours |
| PaO2/FiO2 | Categorical | No ARDS |
| Albumin | Continuous | 4 |
| Lactate | Continuous | 0 |
| Fluid balance | Continuous | 0 |
| Heart rate | Continuous | 80 |
| Temperature | Continuous | 98.6 |
| Bicarbonate | Continuous | 24 |
| Creatinine | Continuous | 1 |
| Blood glucose | Continuous | 160 |
| WBC | Continuous | 8 |
| Potassium | Continuous | 4 |
| Sodium | Continuous | 140 |
| HGB | Continuous | 16 if Male, 14 if Female |
| HCT | Continuous | 46 if Male, 42 if Female |
| PLT | Continuous | 300 |

**Supplemental Table 2.** Mortality prediction variables with definitions

| **Feature** | **Feature Type** | **Number (%) of missingness** | **Definition** |
| --- | --- | --- | --- |
| *Baseline Patient Characteristics* | | | |
| Age | Continuous | 0 (0) | N/A |
| Sex | Categorical | 0 (0) | N/A |
| *ICU Admission Information* | | | |
| Admission diagnosis | Categorical | 0 (0) | Burn, cardiovascular, dermatology, electrolyte abnormalities, endocrine, fever, gastrointestinal, hematologic, hepatic, infection, mental health, neoplasm, neurology, pneumonia, pregnancy, pulmonary, renal, respiratory, respiratory failure, sepsis, shock, syncope, toxicology/ingestion, trauma, weakness, or other |
| ICU Type | Categorical | 0 (0) | Burn, cardiac, cardiothoracic, medical, neurosciences, surgical, or mixed |
| *24 Hours After ICU Admission* | | | |
| *Severity of Illness* | | | |
| APACHE II at 24 hours | Continuous | 0 (0) | N/A |
| SOFA at 24 hours | Continuous | 0 (0) | N/A |
| *Vital Signs* | | | |
| Heart rate at 24 hours | Continuous | 3 (0.3) | Highest heart rate at 24 hours |
| *Ind_miss heart rate at 24 hours* |  |  | Indicator of missingness for heart rate at 24 hours |
| SBP at 24 hours | Categorical | 12 (1.21) | Lowest systolic blood pressure at 24 hours. This variable is categorized into two levels by cut point 90 |
| *Ind_miss SBP at 24 hours* |  |  | Indicator of missingness for SBP at 24 hours. This variable indicates missingness into two categories depending on the level of vasopressor at 24 hours |
| Temperature at 24 hours | Continuous | 26 (2.62) | The most extreme temperature at 24 hours comparing with the normal value, 97.97 |
| *Ind_miss temperature at 24 hours* |  |  | Indicator of missingness for temperature at 24 hours |
| *ARDS Classification* | | | |
| PaO_2_:FiO_2_ at 24 hours | Categorical | 622 (62.76) | The lowest PaO_2_:FiO_2_ at 24 hours. This variable is categorized into four levels by cut points 100, 200, and 300. |
| *Ind_miss PaO_2_:FiO_2_ at 24 hours* |  |  | Indicator of missingness for PaO_2_:FiO_2_ at 24 hours |
| *Supportive Care Devices* | | | |
| CRRT at 24 hours | Categorical | 0 (0) |  |
| Mechanical ventilation at 24 hours | Categorical | 0 (0) | N/A |
| *Serum Laboratory Values* | | | |
| Albumin at 24 hours | Continuous | 521 (52.57) | The lowest albumin at 24 hours |
| *Ind_miss albumin at 24 hours* |  |  | Indicator of missingness for albumin at 24 hours |
| Bicarbonate at 24 hours | Continuous | 46 (4.64) | The most extreme bicarbonate at 24 hours comparing with the normal value, 24 |
| *Ind_miss bicarbonate & creatinine & sodium at 24 hours* |  |  | Indicator of missingness for bicarbonate & creatinine & sodium at 24 hours |
| Blood glucose at 24 hours | Continuous | 31 (3.13) | The most extreme blood glucose at 24 hours comparing with the normal value, 125 |
| *Ind_miss blood glucose at 24 hours* |  |  | Indicator of missingness for blood glucose at 24 hours |
| Creatinine at 24 hours | Continuous | 46 (4.64) | The highest creatinine at 24 hours |
| HCT at 24 hours | Continuous | 80 (8.07) | The lowest HCT at 24 hours |
| HGB at 24 hours | Continuous | 80 (8.07) | The lowest HGB at 24 hours |
| *Ind_miss HGB & HCT at 24 hours* |  |  | Indicator of missingness for HGB & HCT at 24 hours. This variable indicates missingness into two categories depending on the level of Sex |
| Lactate at 24 hours | Continuous | 670 (67.61) | The highest lactate at 24 hours |
| *Ind_miss lactate at 24 hours* |  |  | Indicator of missingness for lactate at 24 hours |
| pH at 24 hours | Categorical | 582 (58.73) | The most extreme pH at 24 hours comparing with the normal value, 7.4. This variable is categorized into three levels by two cut points, 7.2 and 7.5 |
| *Ind_miss pH at 24 hours* |  |  | Indicator of missingness for pH at 24 hours |
| PLT at 24 hours | Continuous | 83 (8.38) | The most extreme PLT at 24 hours comparing with the normal value, 300. |
| *Ind_miss PLT at 24 hours* |  |  | Indicator of missingness for PLT at 24 hours |
| Potassium at 24 hours | Continuous | 44 (4.44) | The most extreme potassium at 24 hours comparing with the normal value, 4.35 |
| *Ind_miss potassium at 24 hours* |  |  | Indicator of missingness for potassium at 24 hours |
| Sodium at 24 hours | Continuous | 46 (4.64) | The most extreme sodium at 24 hours comparing with the normal value, 140 |
| WBC at 24 hours | Continuous | 81 (8.17) | The most extreme WBC at 24 hours comparing with the normal value, 7.75 |
| *Ind_miss WBC at 24 hours* |  |  | Indicator of missingness for WBC at 24 hours |
| Fluid balance at 24 hours | Continuous | 100 (10.09) |  |
| *Ind_miss fluid balance at 24 hours* |  |  | Indicator of missingness for fluid balance at 24 hours |
| *Medication Data at 24 Hours* | | | |
| MRC-ICU at 24 hours | Continuous | 0 (0) | N/A |
| Vasopressor at 24 hours | Categorical | 0 (0) | N/A |
| APACHE: Acute Physiology and Chronic Health Evaluation; CRRT: continuous renal replacement therapy; HCT: hematocrit; HGB: hemoglobin; MRC-ICU: Medication Regimen Complexity – Intensive Care Unit; PaO_2_:FiO_2_: ratio of partial pressure of arterial oxygen to fraction of inspired oxygen; PLT: platelets; SBP: systolic blood pressure; SOFA: Sequential Organ Failure Assessment; WBC: white blood cells | | | |

**Supplemental Table 3.** Baseline study cohort demographics – validation set

|  | All (n=4878) | Mortality (n=964) | No mortality (n=3914) | p-value |
| --- | --- | --- | --- | --- |
| *ICU baseline* | | | | |
| Age, mean (SD) | 58.87 (16.47) | 62.46 (14.53) | 57.98 (16.80) | 0.00 |
| Male sex, n (%) | 2532 (51.91) | 528 (54.77) | 2004 (51.20) | 0.05 |
| *ICU Type, n (%)* | | | | 0.57 |
| Mixed ICU | 2 (0.04) | 0 (0.00) | 2 (0.05) |  |
| Burn ICU | 32 (0.66) | 4 (0.41) | 28 (0.72) |  |
| Cardiac ICU | 41 (0.84) | 12 (1.24) | 29 (0.72) |  |
| Cardiothoracic ICU | 15 (0.31) | 3 (0.31) | 12 (0.31) |  |
| Medical ICU | 4631 (94.94) | 917 (95.12) | 3714 (94.89) |  |
| Neurosciences ICU | 31 (0.64) | 5 (0.52) | 26 (0.66) |  |
| Surgical ICU | 126 (2.58) | 23 (2.39) | 103 (2.63) |  |
| *Primary ICU admission diagnosis, n (%)* | | | | 0.00 |
| Burn | 12 (0.25) | 1 (0.10) | 11 (0.28) |  |
| Cardiovascular | 364 (7.46) | 58 (6.02) | 306 (7.82) |  |
| Dermatology | 34 (0.70) | 5 (0.52) | 29 (0.74) |  |
| Electrolyte Abnormalities | 47 (0.96) | 5 (0.52) | 42 (1.07) |  |
| Endocrine | 200 (4.10) | 8 (0.83) | 192 (4.91) |  |
| Fever | 75 (1.54) | 11 (1.14) | 64 (1.64) |  |
| Gastrointestinal | 573 (11.75) | 76 (7.88) | 497 (12.70) |  |
| Hematologic | 347 (7.11) | 75 (7.78) | 272 (6.95) |  |
| Hepatic | 188 (3.85) | 66 (6.85) | 122 (3.12) |  |
| Infection | 399 (8.18) | 132 (13.69) | 267 (6.82) |  |
| Mental Health | 31 (0.64) | 1 (0.10) | 30 (0.77) |  |
| Neoplasm | 135 (2.77) | 29 (3.01) | 106 (2.71) |  |
| Neurology | 331 (6.79) | 57 (5.91) | 274 (7.00) |  |
| Pneumonia | 148 (3.03) | 34 (3.53) | 114 (2.91) |  |
| Pregnancy | 21 (0.43) | 1 (0.10) | 20 (0.51) |  |
| Pulmonary | 88 (1.8) | 11 (1.14) | 77 (1.97) |  |
| Renal | 180 (3.69) | 33 (3.42) | 147 (3.76) |  |
| Respiratory | 420 (8.61) | 84 (8.71) | 336 (8.58) |  |
| Respiratory failure | 396 (8.12) | 118 (12.24) | 278 (7.10) |  |
| Sepsis | 559 (11.46) | 118 (12.24) | 441 (11.27) |  |
| Shock | 49 (1.00) | 10 (1.04) | 39 (1.00) |  |
| Syncope | 18 (0.37) | 2 (0.21) | 16 (0.41) |  |
| Toxicology/Ingestion | 88 (1.80) | 4 (0.41) | 84 (2.15) |  |
| Trauma | 12 (0.25) | 2 (0.21) | 10 (0.26) |  |
| Weakness | 25 (0.51) | 4 (0.41) | 21 (0.54) |  |
| Other | 138 (2.83) | 19 (1.97) | 119 (3.04) |  |
| *24 h after ICU admission* | | | | |
| *Severity of illness, mean (SD)* | | | | |
| APACHE II Score | 15.92 (6.26) | 18.82 (6.40) | 15.20 (6.02) | 0.00 |
| SOFA Score | 7.00 (4.48) | 10.31 (4.71) | 6.18 (4.03) | 0.00 |
| *Vital Signs* |  |  |  |  |
| Heart rate, mean (SD) | 110.14 (21.77) | 115.92 (24.20) | 108.71 (20.89) | 0.00 |
| Systolic blood pressure abnormal (< 90 mmHg), n (%) | 1305 (31.36) | 285 (41.73) | 1020 (29.32) | 0.00 |
| Temperature (F), mean (SD) | 98.71 (2.69) | 98.53 (4.70) | 98.75 (1.94) | 0.17 |
| *ARDS Classification, n (%)* | | | | 0.00 |
| Mild | 386 (22.13) | 104 (15.76) | 282 (26.01) |  |
| Moderate | 667 (38.25) | 265 (40.15) | 402 (37.08) |  |
| Severe | 408 (23.39) | 227 (34.39) | 181 (16.70) |  |
| *Supportive devices, n (%)* | | | | |
| CRRT at 24 h | 611 (12.53) | 316 (32.78) | 295 (7.54) | 0.00 |
| MV at 24 h | 1415 (29.01) | 504 (52.28) | 911 (23.28) | 0.00 |
| *Serum laboratory values, mean (SD)* | | | | |
| Albumin mg/dL | 2.61 (0.54) | 2.49 (0.54) | 2.65 (0.54) | 0.00 |
| Bicarbonate mEq/L | 22.69 (6.53) | 20.56 (7.52) | 23.21 (6.16) | 0.00 |
| Creatinine mg/dL | 2.03 (2.37) | 2.46 (2.11) | 1.92 (2.42) | 0.00 |
| Glucose mg/dL | 154.35 (93.26) | 165.70 (96.27) | 151.55 (92.31) | 0.00 |
| Lactate mmol/L | 3.33 (4.25) | 5.24 (5.84) | 2.21 (2.31) | 0.00 |
| Potassium mEq/L | 4.12 (0.83) | 4.37 (0.95) | 4.06 (0.79) | 0.00 |
| pH < 7.2, n (%) | 191 (9.49) | 144 (20.06) | 47 (3.63) | 0.00 |
| pH > 7.5, n (%) | 142 (7.05) | 33 (4.60) | 109 (8.42) | 0.00 |
| Sodium mEq/L | 137.13 (6.49) | 137.36 (6.76) | 137.07 (6.42) | 0.25 |
| Hemoglobin g/dL | 9.69 (2.26) | 9.35 (2.32) | 9.77 (2.24) | 0.00 |
| Hematocrit % | 30.21 (7.01) | 29.37 (7.27) | 30.42 (6.93) | 0.00 |
| Platelets x 10^3^/μL | 199.57 (136.92) | 180.76 (146.07) | 204.30 (134.13) | 0.00 |
| White blood cells x 10^3^/μL | 13.44 (18.64) | 17.15 (19.91) | 12.52 (18.19) | 0.00 |
| Fluid balance at 24 h (L), mean (SD) | 1.81 (2.98) | 2.50 (3.25) | 1.64 (2.88) | 0.00 |
| *Medications* | | | | |
| MRC-ICU, mean (SD) | 9.91 (6.64) | 13.75 (7.71) | 8.96 (5.99) | 0.00 |
| Vasopressor at 24 h, n (%) | 2106 (43.17) | 659 (68.36) | 1447 (36.97) | 0.00 |

**Supplemental Table 4.** Data missingness in validation set

| **Feature** | **Number (%) Missingness** |
| --- | --- |
| Age | 0 (0) |
| Sex | 0 (0) |
| MRC-ICU score at 24 hours | 0 (0) |
| ICU type | 0 (0) |
| Mechanical ventilation at 24 hours | 0 (0) |
| Fluid balance at 24 hours | 0 (0) |
| Admission diagnosis | 0 (0) |
| Fluid balance at 24 hours | 4 (0.08) |
| CRRT at 24 hours | 0 (0) |
| Vasopressor at 24 hours | 0 (0) |
| Heart rate at 24 hours | 3 (0.06) |
| SBP at 24 hours | 716 (14.68) |
| Temperature at 24 hours | 499 (10.23) |
| pH at 24 hours | 2865 (58.73) |
| Bicarbonate at 24 hours | 222 (4.55) |
| Creatinine at 24 hours | 192 (3.94) |
| Blood glucose at 24 hours | 124 (2.54) |
| WBC at 24 hours | 295 (6.05) |
| Lactate at 24 hours | 3095 (63.45) |
| Potassium at 24 hours | 192 (3.94) |
| Sodium at 24 hours | 183 (3.75) |
| Albumin at 24 hours | 2033 (41.68) |
| HGB at 24 hours | 269 (5.51) |
| HCT at 24 hours | 276 (5.66) |
| PLT at 24 hours | 293 (6.01) |
| PaO_2_:FiO_2_ at 24 hours | 3134 (64.25) |

**Supplemental Table 5.** AUROC for mortality prediction models on validation set

|  | **AUROC** |
| --- | --- |
| *APACHE II* | 0.66, 0.64-0.68 |
| *SOFA* | 0.75, 0.73-0.77 |
| *MRC-ICU + SOFA + APACHE II* | 0.72, 0.70-0.74 |
| *Linear Logistic* | 0.75, 0.73-0.77 |
| *Nature Cubic Splines Logistic* | 0.69, 0.67-0.71 |
| *Smoothing Splines Logistic* | 0.73, 0.71-0.75 |
| *Local Linear Logistic* | 0.73, 0.71-0.75 |
| *Random Forest* | 0.78, 0.76-0.8 |
| *SVM* | 0.74, 0.72-0.76 |
| *XGBoost* | 0.73, 0.71-0.75 |
| AUROC: area under the receiver operating characteristic | |

**Supplemental Table 6.** Accuracy, sensitivity, specificity, negative predictive value, and positive predictive value for mortality prediction on validation set

| **Maximizing INF** | **Accuracy** | **Sensitivity** | **Specificity** | **PPV** | **NPV** |
| --- | --- | --- | --- | --- | --- |
| *APACHE II* | 0.67, 0.65-0.68 | 0.49, 0.45-0.52 | 0.71, 0.70-0.73 | 0.29, 0.27-0.32 | 0.85, 0.84-0.86 |
| *SOFA* | 0.56, 0.55-0.58 | 0.83, 0.80-0.85 | 0.50, 0.48-0.52 | 0.29, 0.27-0.31 | 0.92, 0.91-0.93 |
| *MRC-ICU + SOFA + APACHE II* | 0.67, 0.65-0.68 | 0.65, 0.62-0.68 | 0.67, 0.66-0.69 | 0.33, 0.31-0.35 | 0.89, 0.87-0.90 |
| *Linear Logistic* | 0.66, 0.64-0.67 | 0.71, 0.68-0.74 | 0.64, 0.63-0.66 | 0.33, 0.31-0.35 | 0.90, 0.89-0.91 |
| *Nature Cubic Splines Logistic* | 0.63, 0.61-0.64 | 0.65, 0.62-0.68 | 0.62, 0.61-0.64 | 0.30, 0.28-0.32 | 0.88, 0.87-0.89 |
| *Smoothing Splines Logistic* | 0.67, 0.66-0.68 | 0.67, 0.64-0.70 | 0.67, 0.65-0.68 | 0.33, 0.31-0.35 | 0.89, 0.88-0.90 |
| *Local Linear Logistic* | 0.68, 0.66-0.69 | 0.65, 0.62-0.68 | 0.68, 0.67-0.70 | 0.34, 0.31-0.36 | 0.89, 0.88-0.90 |
| *Random Forest* | 0.55, 0.53-0.56 | 0.88, 0.86-0.90 | 0.46, 0.45-0.48 | 0.29, 0.27-0.30 | 0.94, 0.93-0.95 |
| *SVM* | 0.66, 0.64-0.67 | 0.72, 0.69-0.75 | 0.64, 0.63-0.66 | 0.33, 0.31-0.35 | 0.90, 0.89-0.91 |
| *XGBoost* | 0.74, 0.73-0.75 | 0.50, 0.47-0.53 | 0.80, 0.79-0.81 | 0.38, 0.36-0.41 | 0.87, 0.86-0.88 |
| **Maximizing MCC** | **Accuracy** | **Sensitivity** | **Specificity** | **PPV** | **NPV** |
| *APACHE II* | 0.67, 0.65-0.68 | 0.49, 0.45-0.52 | 0.71, 0.70-0.73 | 0.29, 0.27-0.32 | 0.85, 0.84-0.86 |
| *SOFA* | 0.75, 0.73-0.76 | 0.56, 0.53-0.59 | 0.79, 0.78-0.81 | 0.40, 0.37-0.43 | 0.88, 0.87-0.89 |
| *MRC-ICU + SOFA + APACHE II* | 0.68, 0.66-0.69 | 0.64, 0.61-0.67 | 0.69, 0.67-0.70 | 0.33, 0.31-0.36 | 0.89, 0.87-0.90 |
| *Linear Logistic* | 0.71, 0.70-0.72 | 0.64, 0.61-0.67 | 0.72, 0.71-0.74 | 0.36, 0.34-0.39 | 0.89, 0.88-0.90 |
| *Nature Cubic Splines Logistic* | 0.63, 0.61-0.64 | 0.65, 0.62-0.68 | 0.62, 0.61-0.64 | 0.30, 0.28-0.32 | 0.88, 0.87-0.89 |
| *Smoothing Splines Logistic* | 0.75, 0.74-0.76 | 0.49, 0.46-0.53 | 0.81, 0.80-0.83 | 0.39, 0.37-0.42 | 0.87, 0.86-0.88 |
| *Local Linear Logistic* | 0.75, 0.74-0.77 | 0.47, 0.44-0.51 | 0.82, 0.81-0.83 | 0.40, 0.37-0.43 | 0.86, 0.85-0.87 |
| *Random Forest* | 0.67, 0.66-0.69 | 0.75, 0.72-0.78 | 0.65, 0.64-0.67 | 0.35, 0.33-0.37 | 0.91, 0.90-0.92 |
| *SVM* | 0.77, 0.75-0.78 | 0.49, 0.45-0.52 | 0.83, 0.82-0.85 | 0.42, 0.39-0.45 | 0.87, 0.86-0.88 |
| *XGBoost* | 0.78, 0.76-0.79 | 0.36, 0.33-0.39 | 0.88, 0.87-0.89 | 0.42, 0.39-0.46 | 0.85, 0.84-0.86 |
| **Maximizing F1** | **Accuracy** | **Sensitivity** | **Specificity** | **PPV** | **NPV** |
| *APACHE II* | 0.67, 0.65-0.68 | 0.49, 0.45-0.52 | 0.71, 0.70-0.73 | 0.29, 0.27-0.32 | 0.85, 0.84-0.86 |
| *SOFA* | 0.75, 0.73-0.76 | 0.56, 0.53-0.59 | 0.79, 0.78-0.81 | 0.40, 0.37-0.43 | 0.88, 0.87-0.89 |
| *MRC-ICU + SOFA + APACHE II* | 0.69, 0.68-0.70 | 0.61, 0.58-0.64 | 0.71, 0.69-0.72 | 0.34, 0.32-0.36 | 0.88, 0.87-0.89 |
| *Linear Logistic* | 0.71, 0.70-0.73 | 0.63, 0.60-0.66 | 0.74, 0.72-0.75 | 0.37, 0.35-0.39 | 0.89, 0.88-0.90 |
| *Nature Cubic Splines Logistic* | 0.63, 0.62-0.65 | 0.64, 0.61-0.67 | 0.63, 0.62-0.65 | 0.30, 0.28-0.32 | 0.88, 0.86-0.89 |
| *Smoothing Splines Logistic* | 0.75, 0.74-0.76 | 0.49, 0.46-0.53 | 0.81, 0.80-0.83 | 0.39, 0.37-0.42 | 0.87, 0.86-0.88 |
| *Local Linear Logistic* | 0.75, 0.74-0.77 | 0.47, 0.44-0.51 | 0.82, 0.81-0.83 | 0.40, 0.37-0.43 | 0.86, 0.85-0.87 |
| *Random Forest* | 0.77, 0.76-0.78 | 0.59, 0.55-0.62 | 0.81, 0.80-0.82 | 0.44, 0.41-0.46 | 0.89, 0.88-0.90 |
| *SVM* | 0.77, 0.75-0.78 | 0.49, 0.45-0.52 | 0.83, 0.82-0.85 | 0.42, 0.39-0.45 | 0.87, 0.86-0.88 |
| *XGBoost* | 0.78, 0.76-0.79 | 0.36, 0.33-0.39 | 0.88, 0.87-0.89 | 0.42, 0.39-0.46 | 0.85, 0.84-0.86 |
| PPV: positive predictive value; NPV: negative predictive value | | | | | |

**Supplemental Figure 1.** Consort diagram

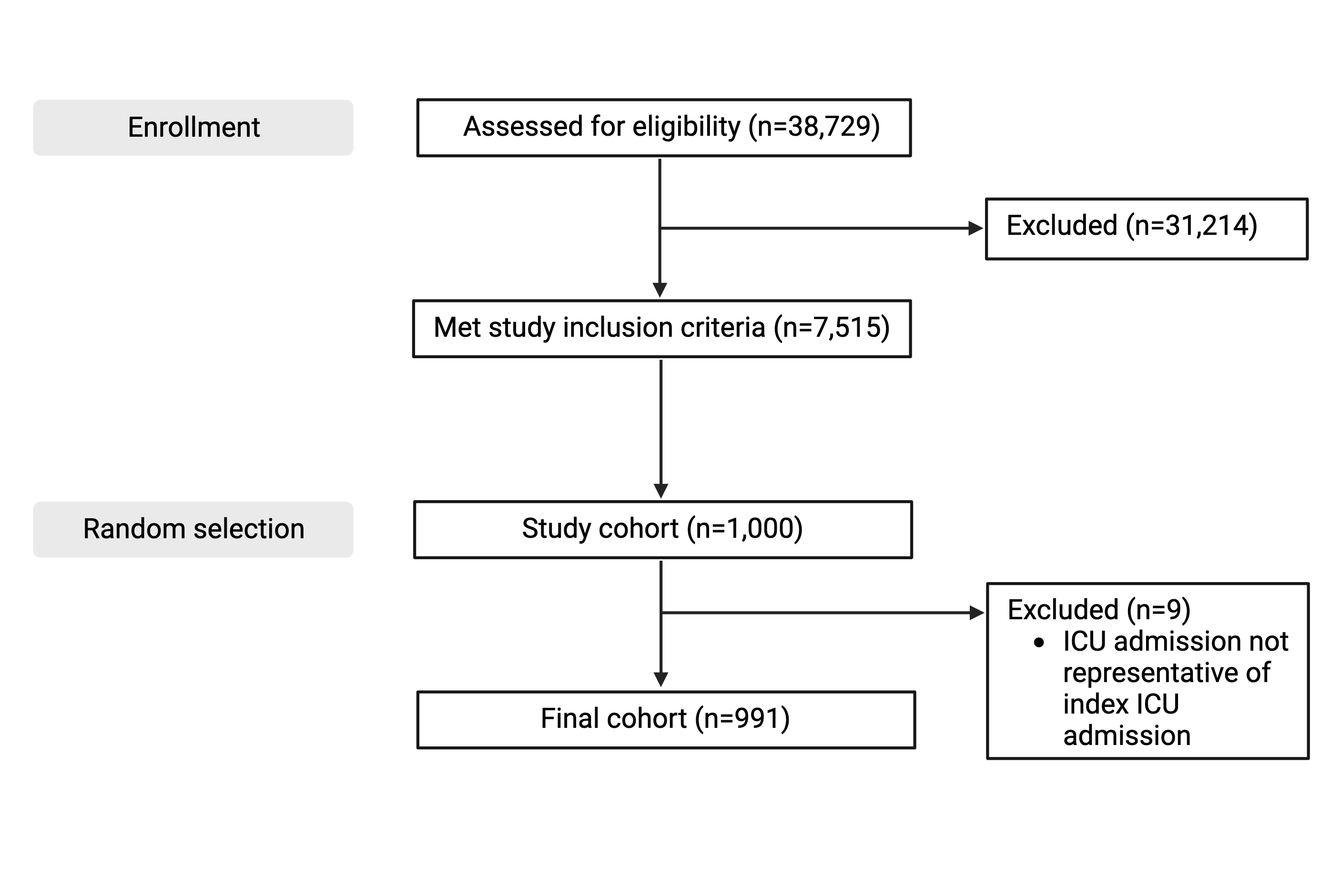

**Supplemental Figure 2.** Feature importance graph for Random Forest

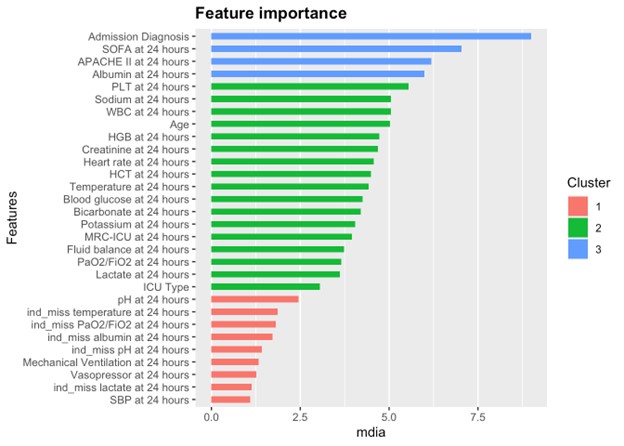

**Supplemental Figure 3.** Feature importance graph for SVM

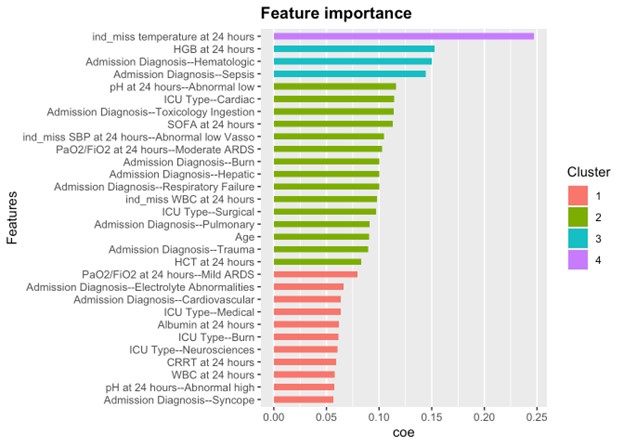

**Supplemental Figure 4.** AUROCs for hospital mortality prediction on validation set

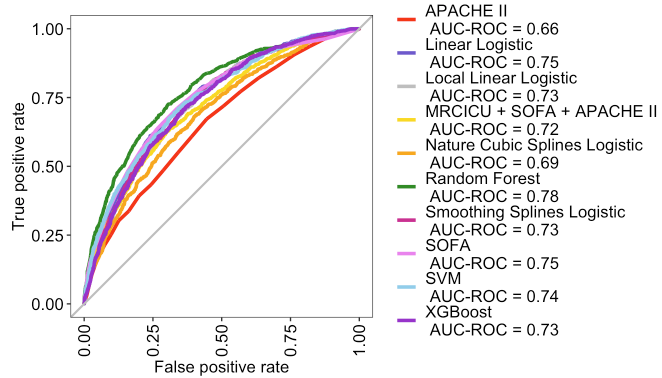
